# Rare variant contribution to cholestatic liver disease in a South Asian population in the United Kingdom

**DOI:** 10.1101/2022.05.05.22274722

**Authors:** Julia Zöllner, Sarah Finer, Kenneth J. Linton, Genes and Health Research Team, David A. van Heel, Catherine Williamson, Peter H. Dixon

## Abstract

**Objectives:** This study assessed the contribution of five genes previously known to be involved in cholestatic liver disease in British Bangladeshi and Pakistani people in the United Kingdom as they are an understudied genetic ancestry group with disproportionate disease burden.

**Methods:** Five genes (*ABCB4, ABCB11, ATP8B1, NR1H4, TJP2*) were interrogated by low/mid whole exome sequencing data of 5236 volunteers. Included were non-synonymous or loss of function (LoF) variants with a minor allele frequency <5%. Variants were filtered and annotated. Rare variant burden analysis was conducted. Variants associated with a phenotype or predicted to be likely pathogenic (LP) underwent protein structure and modelling analysis in silico.

**Results:** Out of 314 non-synonymous variants, 180 fulfilled the inclusion criteria and were mostly heterozygous unless specified. 90 were novel and unique to this cohort and not previously reported in the GnomAD database. Of those novel variants, 22 were considered LP and 9 pathogenic. We identified variants in volunteers with gallstone disease (n=31), intrahepatic cholestasis of pregnancy (ICP, n=16), cholangiocarcinoma and cirrhosis (n=2). Fourteen novel LoF variants were identified: 7 frameshift, 5 introduction of premature stop codon and 2 splice acceptor variants. The rare variant burden was significantly increased in *ABCB11*. A total of 73 variants were assessed for impact at the protein level. Protein modelling demonstrated variants that appeared to likely cause significant structural damage.

**Conclusions:** This study highlights the significant genetic burden contributing to cholestatic liver disease. Novel likely pathogenic and pathogenic variants were identified addressing the underrepresentation of diverse ancestry groups in genomic research.

**WHAT IS KNOWN:** Cholestatic liver diseases encompass a broad range of conditions.

Intrahepatic cholestasis of pregnancy (ICP) is the commonest gestational liver disease.

Genetic and environmental factors contribute to the aetiology of cholestatic disease.

South Asian populations are disproportionally affected.

**WHAT IS NEW HERE:** Exome sequencing analysis in a British Pakistani and Bangladeshi population discovered new genetic mutations.

Pathogenic variants were identified that increase risk of cholestatic liver disease.

Novel variants that contribute to ICP were identified.

## Introduction

Cholestatic liver disease encompasses a broad range of diagnoses that can present with fatigue, pruritus or jaundice[1]. Several genes are implicated, including the *ABCB4* gene coding for the canalicular phosphatidylcholine floppase ABCB4, and the *ABCB11* gene coding for the bile salt export pump (BSEP). Homozygous mutations in *ABCB4* and *ABCB11* cause a spectrum of disease from mild cholestasis to severe progressive familial intrahepatic cholestasis (PFIC), PFIC3 and PFIC2 respectively[2, 3]. *ABCB4* variants also increase the risk of developing drug-induced intrahepatic cholestasis, gallstone disease, gallbladder and bile duct carcinoma, liver cirrhosis and abnormal liver function tests[4]. Other canalicular transporters and their regulators are implicated in the pathogenesis of cholestatic liver disease e.g., *ATP8B1* (a P-type ATPase that flips phospholipids into the cytoplasmic leaflet of the membrane) [3], *NR1H4* (farnesoid X receptor (FXR)) gene[5], and *TJP2* (tight junction protein 2)[6]. While homozygous mutations of these genes are implicated in rare cases of severe familial cholestasis, the evidence base for a role of heterozygous mutations in milder forms of liver disease is limited. *ABCB4* and *ABCB11* are involved in 20% of patients with severe intrahepatic cholestasis of pregnancy (ICP). Heterozygous *ABCB4* variants have also been reported in ICP[7–13]. ICP is the commonest gestational liver disease[14] and may be complicated by preterm birth, meconium-stained amniotic fluid and stillbirth, in association with maternal serum bile acid concentrations ≥40µmol[15, 16]. In Europeans the incidence is 0.62% versus 1.24% in women of Indian and 1.46% of Pakistani origin[17]. Despite the increased prevalence of ICP and other liver conditions like non-alcoholic fatty liver disease in South Asian populations they often remain undiagnosed and under-investigated[18, 19].

Genes and Health is a long-term population-based cohort that assesses the health and disease in British Bangladeshi and British Pakistani people. Using the Genes & Health genomics (whole exome sequencing (WES)) and data linkage to electronic health records (EHR)[20], this study investigated rare variation in a unique British Bangladeshi and Pakistani cohort around 5 candidate loci (*ABCB4, ABCB11, ATP8B1, NR1H4*, and *TJP2*) implicated in cholestatic liver disease.

## Methods

### Study population

Genes & Health has previously been described by Finer et al.[20]. Ethical approval for the study was provided by the South East London National Research Ethics Committee (14/LO/1240) including consent for publishing http://www.genesandhealth.org/volunteer-information)[22]. An individual application to support data access for this study was granted by Genes & Health (reference S00037).

Exome sequencing samples from 5236 Genes & Health volunteers reporting parental relatedness were available for analysis in variant call format files. For the initial analysis a genotype to phenotype approach was employed interrogating 5 gene candidate loci (Table 1). For the rare burden analysis female volunteers without ICP served as controls (n=3048). In a secondary analysis, a phenotype to genotype analysis was used to validate these findings, using ICP as the exemplar. For this approach, electronic health records allowed total serum bile acid concentrations ≥10 µmol to be retrieved from a network of acute hospitals that provide maternity care to (n=15,500) women per year living in east London to identify patients with a diagnosis of liver disease in pregnancy (ICD 10 diagnosis code O26.6), see Supplementary Figure 1. Maternal health records were screened by an experienced clinician to verify a diagnosis of ICP.

**Table 1.** Description of the five gene candidates.

| **Genes** | **Chr** | **Gene product** | **Omim ID** | **Exons** | **Length** | **Associated disease** |
| --- | --- | --- | --- | --- | --- | --- |
| ABCB4 | 7 (7q21.1) | . | 171060 | 27 | 81kb | PFIC-3 |
| ABCB11 | 2 (2q31.1) | BSEP | 603201 | 28 | 115kb | PFIC-2 |
| ATP8B1 | 18 (18q21.31) | FIC1 | 602397 | 28 | 157kb | PFIC-1 |
| TJP2 | 9 (9q21.11) | . | 607709 | 25 | 153kb | PFIC-4 |
| NR1H4 | 12 (12q23.1) | FXR | 603826 | 11 | 90kb | PFIC-5 |

Footnote: Abbreviations: Chr, chromosomes.

### Exome sequencing & bioinformatic pipeline

Low/mid exome sequencing was performed as previously described[21]. The exome sequencing data is being held under a data access agreement at the European Genotype-phenome Archive (www.ebi.ac.uk/ega) under accession numbers EGAD00001005469. Minor allele frequency (MAF) was set at < 5% to include rare and low-frequency genetic variants to allow for a comprehensive evaluation.

### Variant annotation

All protein-altering missense, non-sense, frameshift indels or splice site variants identified in the candidate gene set underwent the same processing as described below. Synonymous variants were excluded from further analysis. Variants were filtered and annotated if they met any of the following inclusion criteria (MAF < 5%): 1. associated with a phenotype; 2. known in the literature; 3. no recorded GnomAD allele frequency; 4. predicted to be likely pathogenic (LP) based on all 7 *in-silico* predictors. To assess the likelihood of functional impact of variants a variety of *in-silico* tools were employed, including Polyphen[22], SIFT[23], CADD[24], Revel[25], MetaLR[26], MetaSVM[26] and M_CAP[27]. Open-source databases (Leiden Open Variation Database – LoVD, and ClinVar) including a commercial database (Mastermind) were interrogated to assess whether variants were reported previously in the literature. The American College of Medical Genetics/Association for Molecular Pathology (ACMG/AMP) guidelines for rare variant interpretation was used to assess variant pathogenicity[28].

### Rare variant burden analysis

To assess the significance of any rare variant burden of SNPs in all 5 gene candidates the exactCMC function in RVTESTS[29] was used. The burden was calculated as the proportion of all ICP cases versus control in the G&H cohort and who had at least one alternate allele. Variants with an allele frequency of 0.01 or less were included. ICP cases (n=18) from the Genomics England database (Project ID 747) – a predominantly European genetic cohort - were accessed to serve as a direct comparison to rare variant burden in G&H.

### Protein structure and modelling analysis

Inclusion criteria for further protein structure and modelling analysis required variants to have an associated phenotype, and/or be predicted to be pathogenic based on all 7 *in-silico* tools. VarMap was used to assess protein sequence variants[30]. 2D representations were designed using the open source tool TOPO2[31]. Variants that were LP or associated with a phenotype underwent further analysis using: (1) Dynamut[32] (2) CUPSAT[33] (3) SNPMuSiC[34]. 3D structural representations were generated using PyMOL software using *ABCB4* with phosphatidylcholine (PDB ID: 7NIV)[35], *ABCB11* open structure (PDB ID: 6LR0)[36], *ATP8B1* (PDB ID: 7PY4)[37], and *NR1H4* (PDB ID: 1OSH)[38]. There is no 3D protein structure available for *TJP2*.

## Results

### Genotype to phenotype analysis

Screening of the 5 candidate genes identified 300 variants; and 180 (166 non-synonymous and 14 loss of function (LoF) variants), were included in the analysis (Figure 1). Most variants identified were heterozygous unless otherwise specified. Further variant interpretation including scoring details of individual *in-silico* predictors, the impact of the coding substitution on disease propensity and conservation information for all variants are presented in Supplementary Table 1.

**Figure 1.**
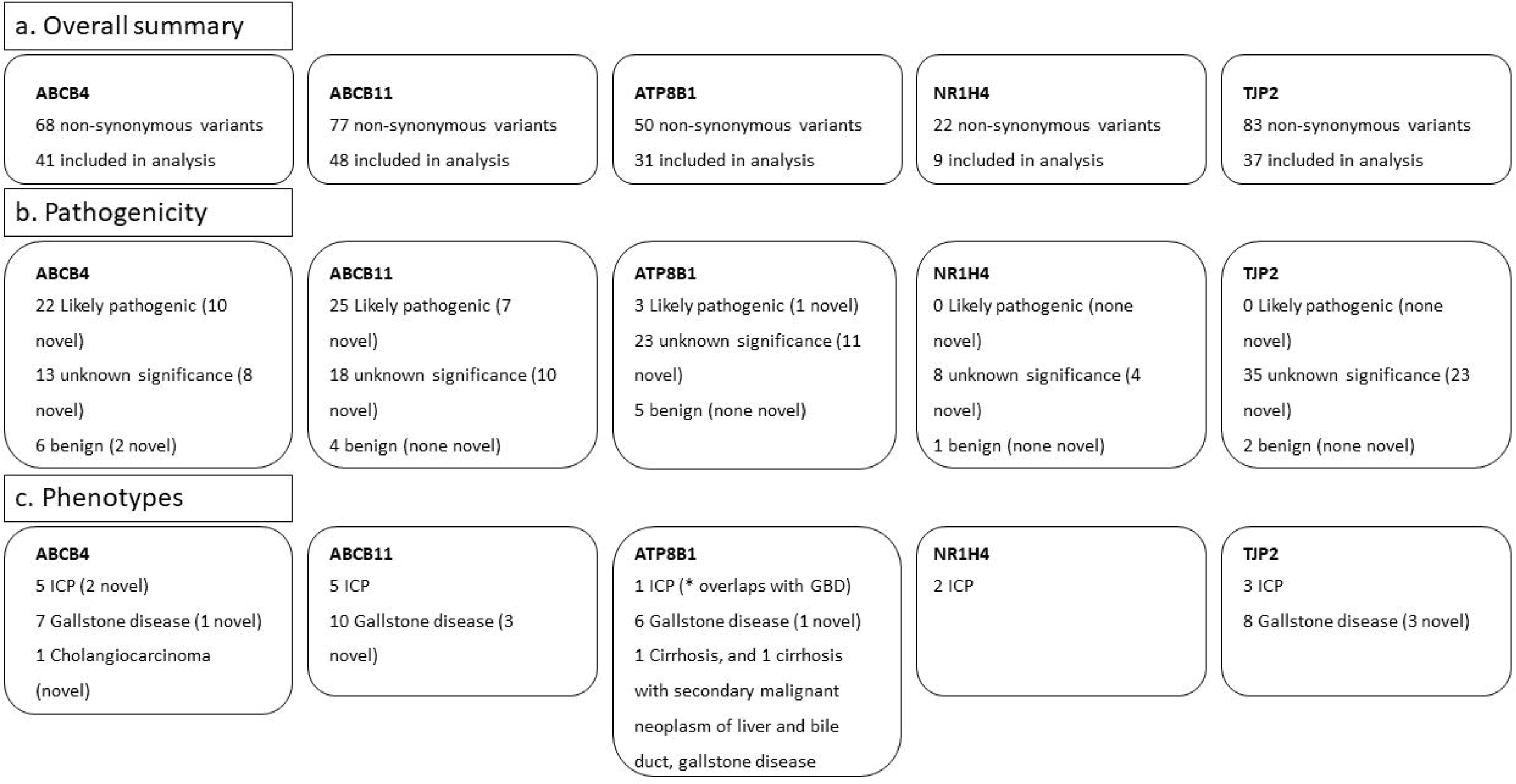
Overall summary of mutational burden discovered in the Genes and Health cohort for all five gene candidates. a. Overall summary, describes all non-synonymous variants identified; b. Pathogenicity, describes breakdown of pathogenicity; c. Phenotypes, describes phenotypes identified. Inclusion criteria (<5% MAF), non-synonymous and at least any of the following: 1. Include all variants with a phenotype 2. Include all variants known in the literature 3. Include all variants with no GnomAD allele frequency but ELGH allele frequency 4. Include all variants with *in-silico* prediction of 7

### Phenotype to genotype analysis

We were able to validate variants of interest in 15 cases reporting ICP, most of whom had documented raised BA concentrations (Table 2). Further, variants discovered in 10 cases with raised BA but an uncertain or no diagnosis of ICP based on their EHR (Supplementary Table 2). This secondary analysis missed 2 individuals from the primary analysis with a diagnosis of ICP as there were no bile acid concentrations available for them. This analysis demonstrated a pragmatic approach to identifying disease causing variants and demonstrates the value of large genomic cohorts linked to electronic health data records.

**Table 2.** Variants identified and TSBA concentration in volunteers with a diagnosis of ICP based on electronic health records.

| All variants in volunteers with raised BA and diagnosis of ICP |  |  |  |  |  |
| --- | --- | --- | --- | --- | --- |
| Volunteers | Gene | Variants | Zygosity | Type | Highest BA (umol/L) |
| 3 | ABCB4 | G1254S / G1261S | het | Non-synonymous | 80.1 |
| 3 | ABCB11 | N591S | het | Non-synonymous |  |
| 4 | ABCB4 | P1050S | het | Non-synonymous | 53 |
| 5 | TJP2 | Q105K | hom | Non-synonymous | 15 |
| 6 | ABCB11 | N591S | het | Non-synonymous | 17 |
| 13 | TJP2 | Q105K | het | Non-synonymous | 25 |
| 15 | ATP8B1 | R384H | het | Non-synonymous | 31.6 |
| 15 | TJP2 | Q105K | hom | Non-synonymous |  |
| 18 | ABCB11 | N591S | het | Non-synonymous | 115 |
| 18 | TJP2 | R21H | het | Non-synonymous |  |
| 20 | ABCB11 | N591S | het | Non-synonymous | 18 |
| 20 | NH1R4 | M173T | het | Non-synonymous |  |
| 21 | ABCB11 | M677V | het | Non-synonymous | 14 |
| 21 | NH1R4 | N358H | het | Non-synonymous |  |
| 22 | ABCB11 | N591S | het | Non-synonymous | 14 |
| 25 | ABCB4 | S99x | het | frameshift | 55 |
| 26 | ABCB11 | V284A | het | Non-synonymous | 35 |
| 26 | ABCB4 | N510S | het | Non-synonymous |  |
| 31 | ABCB4 | T175A | het | Non-synonymous | 32 |
| 32 | ABCB11 | D1284N | het | Non-synonymous | . |
| 32 | ABCB11 | N591S | het | Non-synonymous |  |
| 38 | ABCB11 | N591S | het | Non-synonymous | 45.2 |

**Foot note:** Abbreviations: TSBA, total serum bile acid concentrations; ICP, intrahepatic cholestasis of pregnancy.

### Rare variant burden analysis

We observed in cases versus controls a significant enrichment of rare variants in *ABCB11* (p=0.00247). There was no enrichment in *ABCB4* (p=0.39138), *ATP8B1* (p=0.57957), *TJP2* (p=0.17390), or *NR1H4* (p=0.70232). A further Fisher’s exact analysis compared percentage of rare variants in ICP cases in G&H compared to Genomics England demonstrating that the rare genetic burden was significantly increased in tier 1 gene candidates in British Pakistani and Bangladeshi (*ABCB4*, p=0.0191; *ABCB11*, p=0.0191) but not in *ATP8B1* (p= 0.4857) *NR1H4* (p=0.2286) or *TJP2* (p=0.1039).

### ABCB4 variants

There was a total of 68 *ABCB4* variants identified (Figures 1 and 2). 41 variants fulfilled the inclusion criteria. Variants were identified in people with cholestatic liver disease phenotypes: ICP (n=5 out of 88 women in the analysis), gallstone disease (n=7), and cholangiocarcinoma (n=1) (Table 3). For some identified variants a known cholestatic phenotype had previously been reported in the literature (n=9) whilst others had no phenotype reported (n=19) (Supplementary Table 3). We identified novel LoF variants (n=4): three frameshift (one associated with an ICP phenotype) and one introduction of a premature stop codon (no associated phenotype) (Table 4).

**Table 3.**
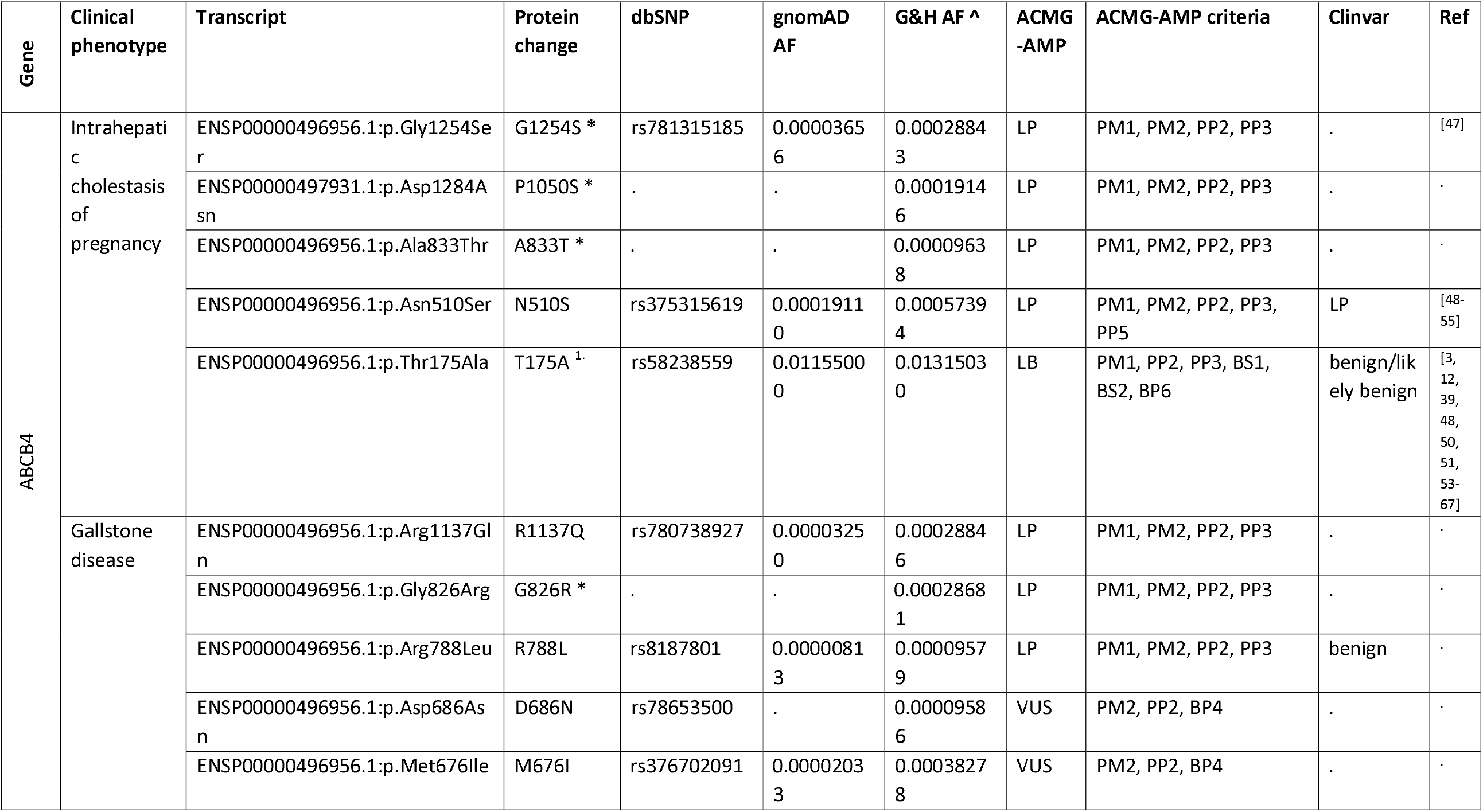
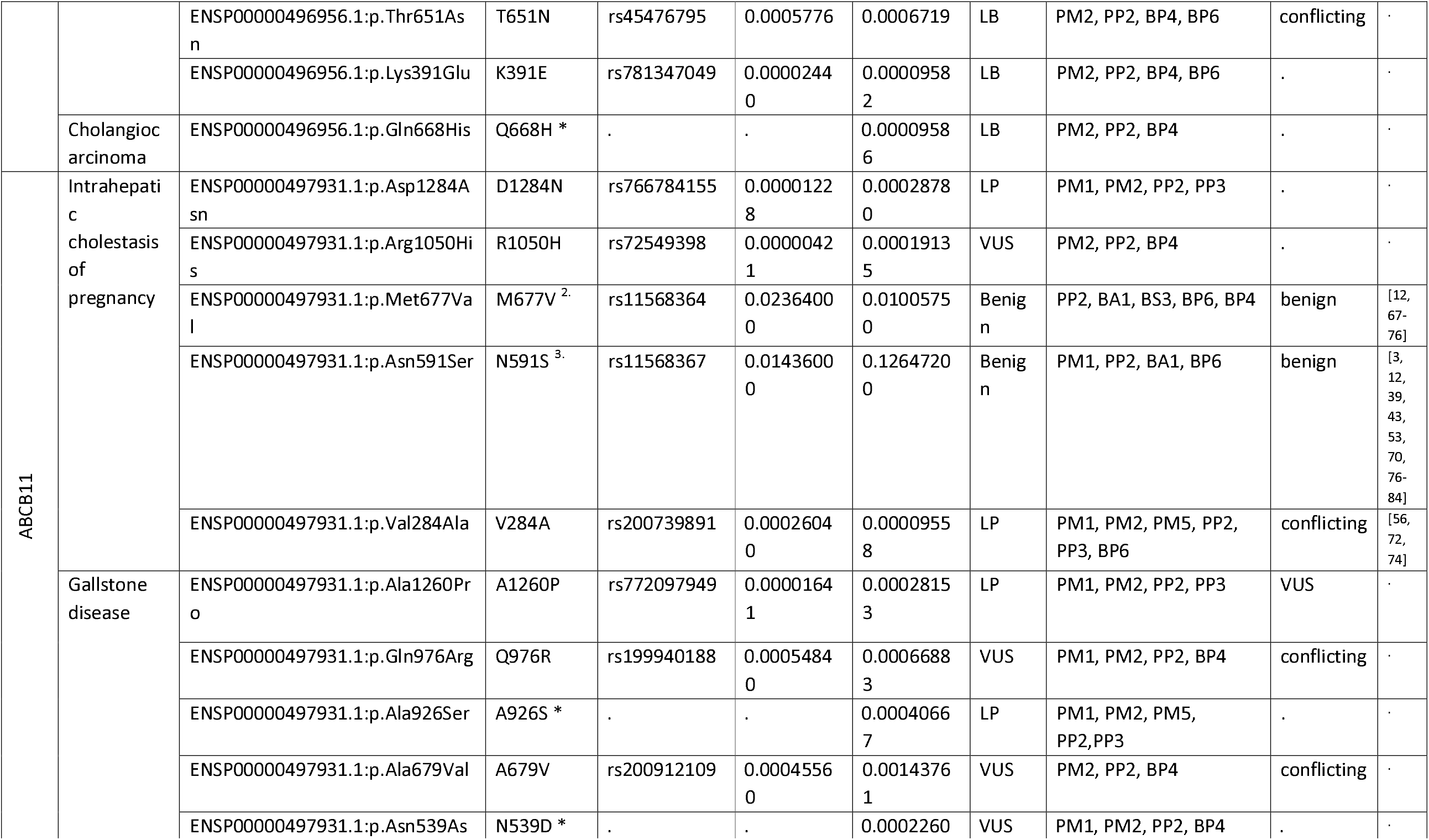
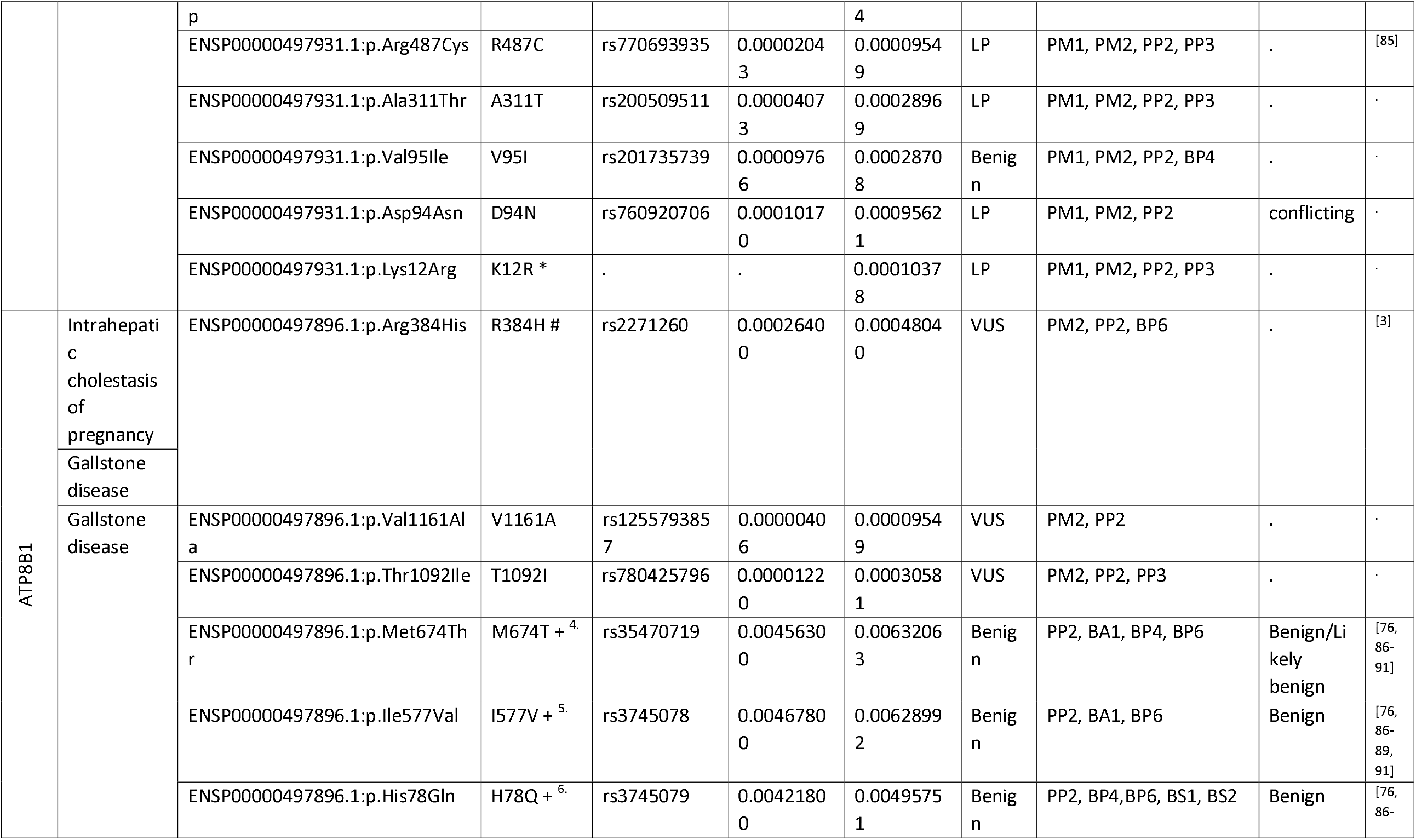
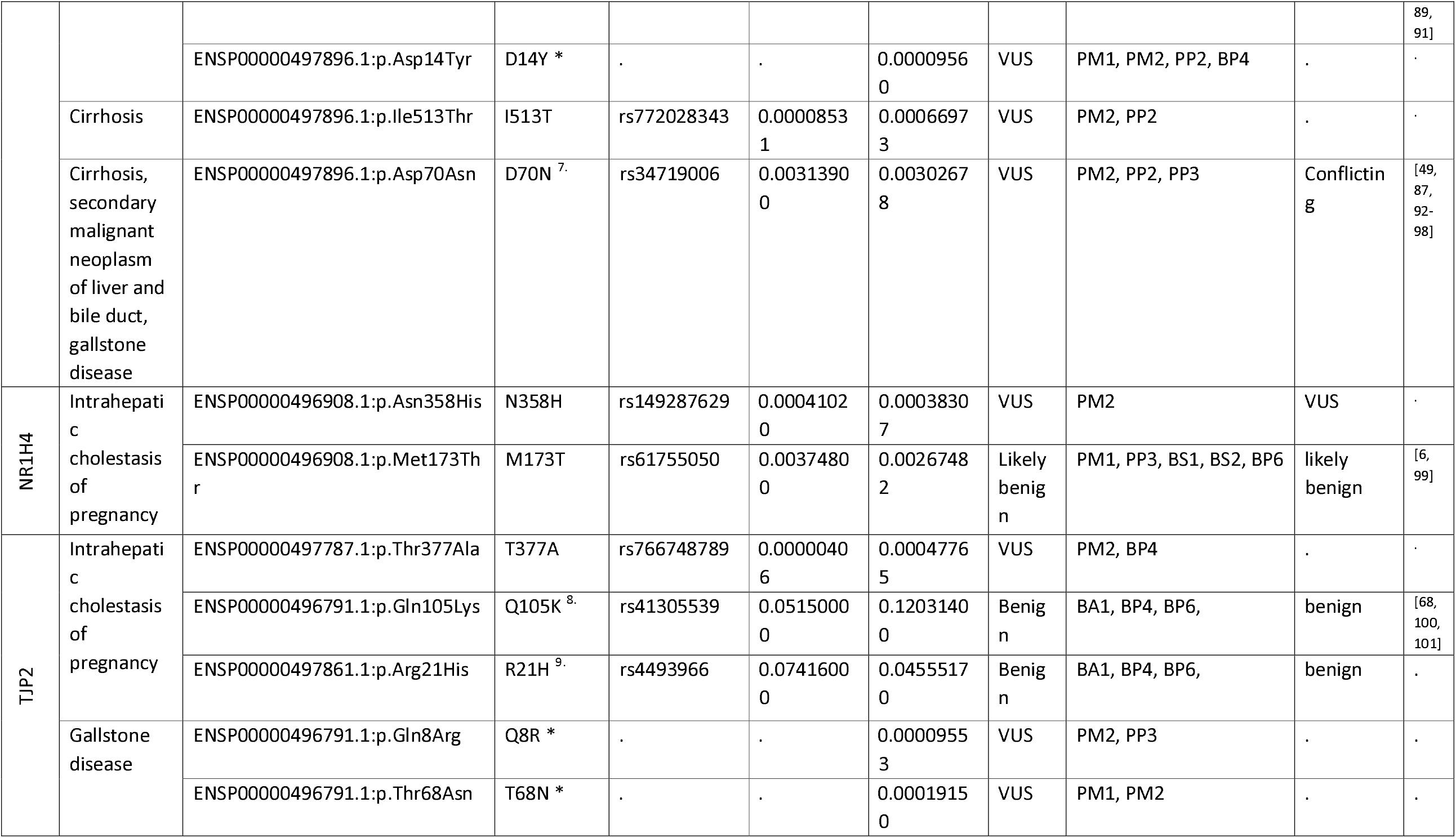
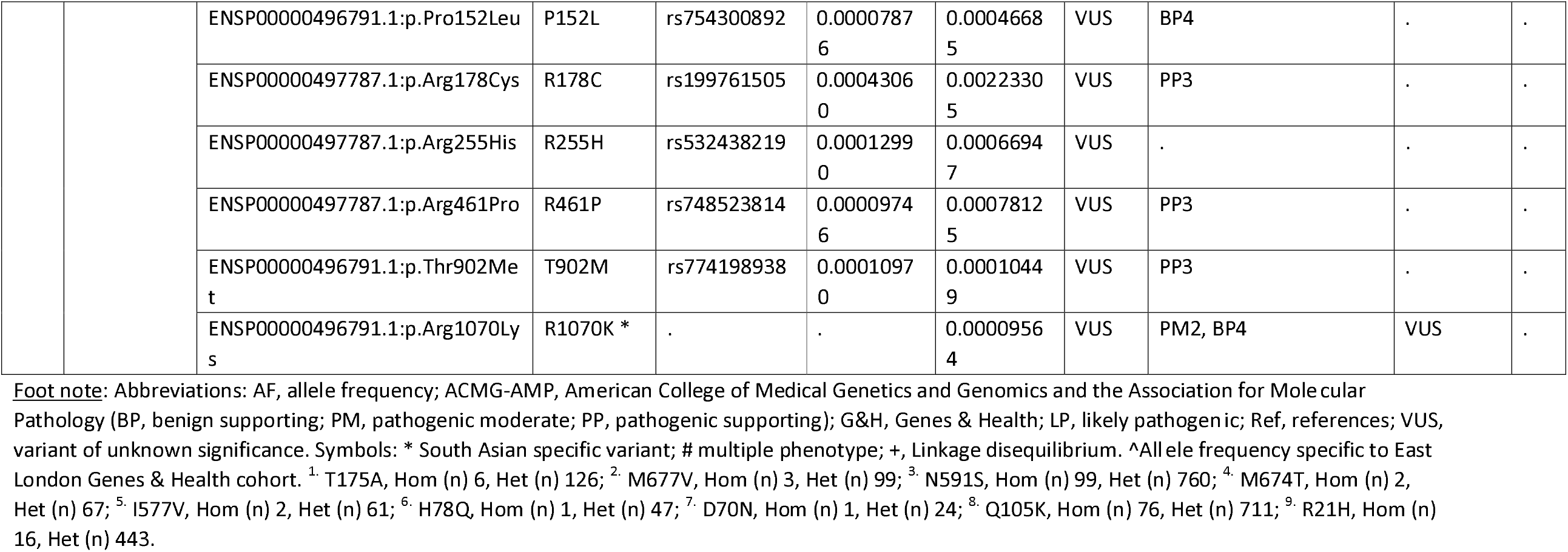
Non-synonymous variants identified in the five gene candidates associated with a cholestatic phenotype in the Genes and Health cohort.

**Table 4.**
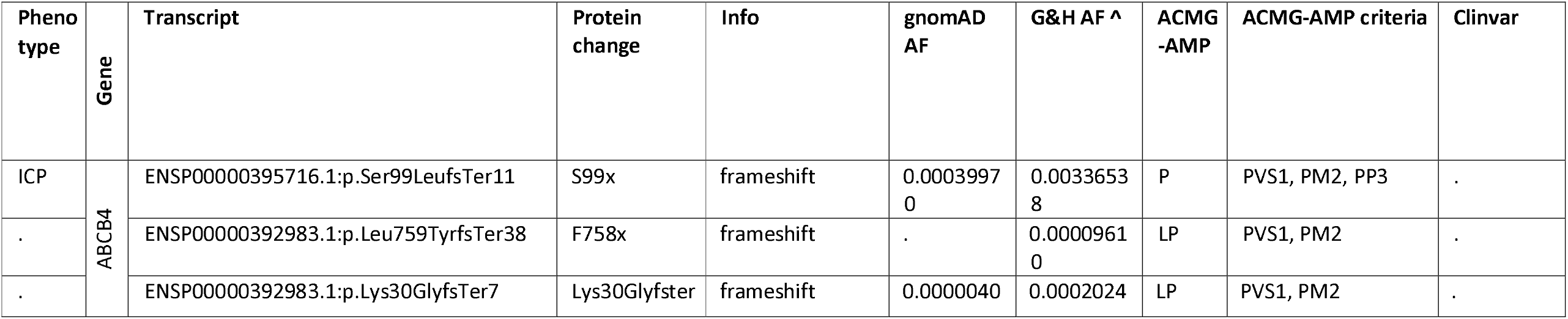
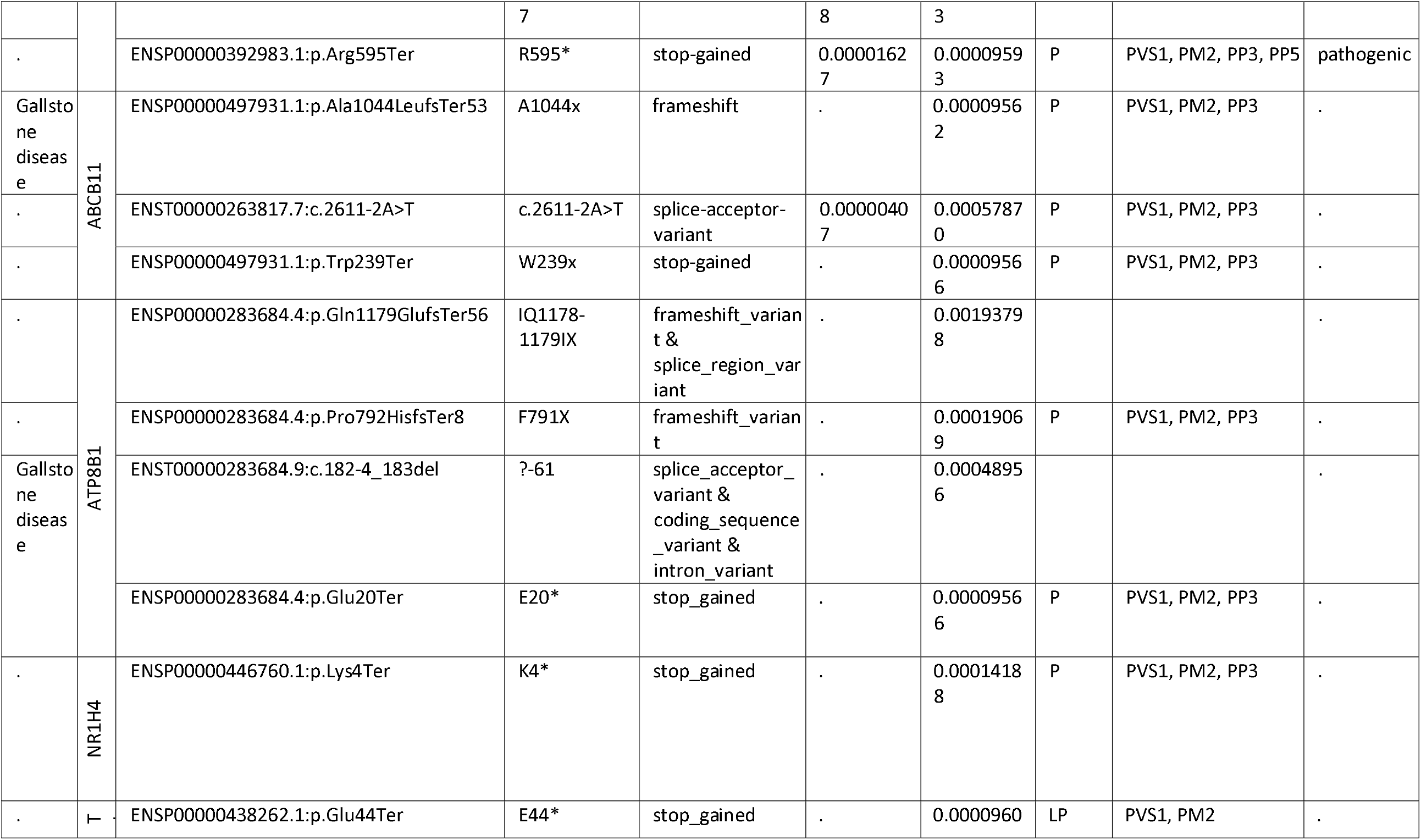
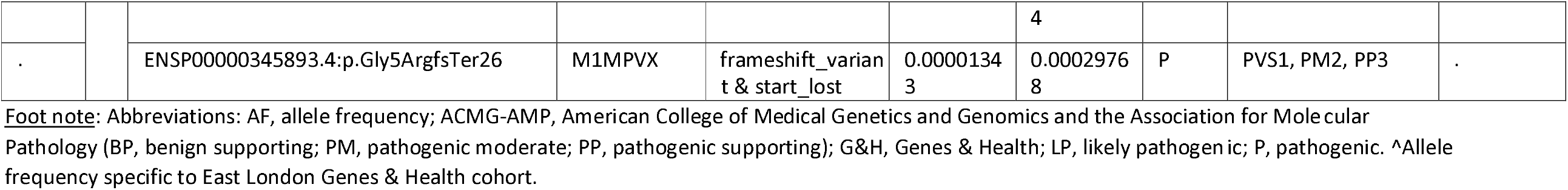
Loss of function variants identified in the five gene candidates.

**Figure 2.**
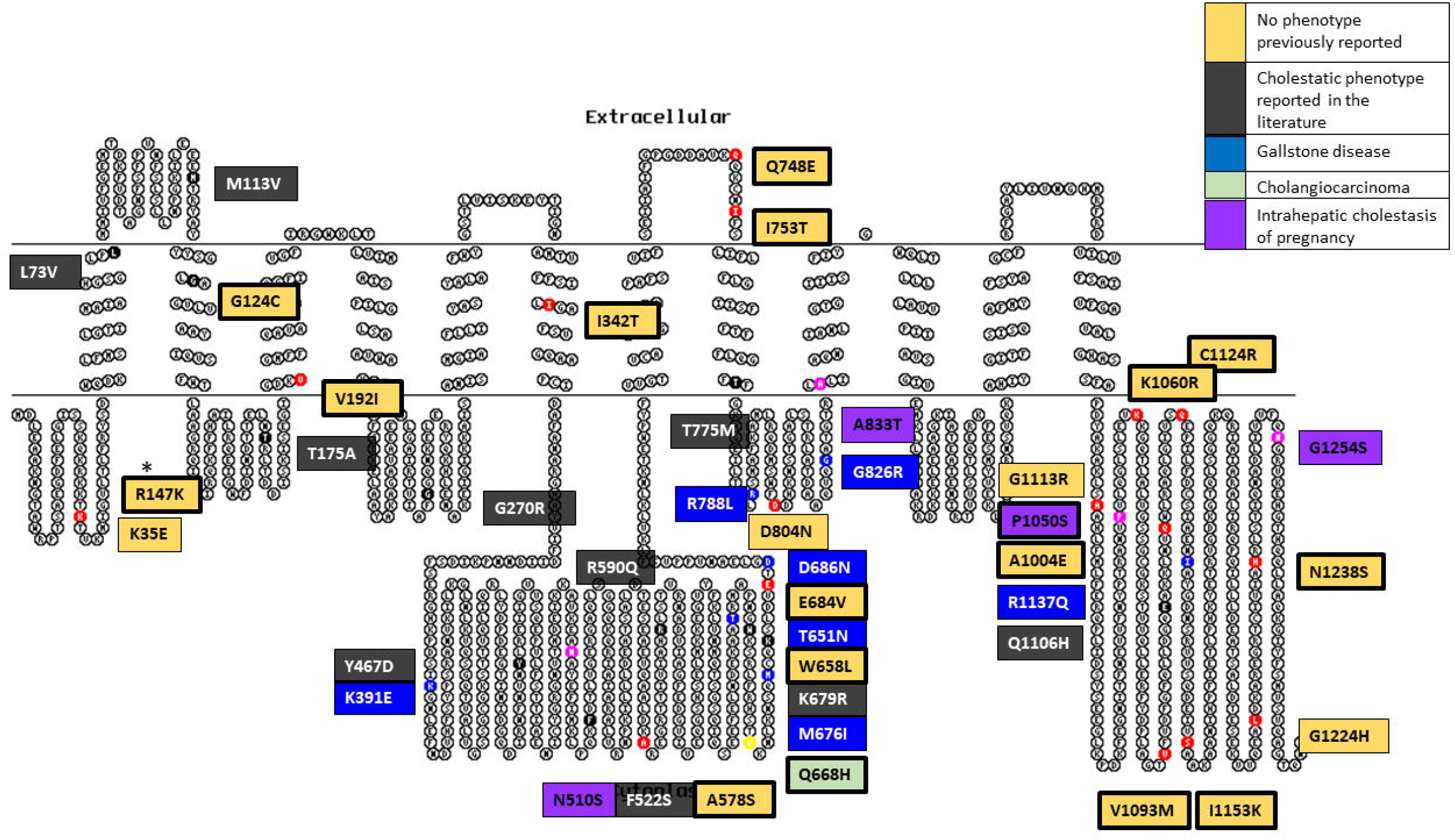
ABCB4 variant summary in a 2-dimensional illustration. Variants are divided into their phenotypic presentation: No phenotype previously reported, cholestatic phenotype reported in the literature, gallstone disease, cholangiocarcinoma, and intrahepatic cholestasis of pregnancy. Bold border represents variants that are unique to the Genes & Health cohort.

### ABCB11 variants

There were 77 *ABCB11* variants identified of which 48 were included in the analysis (Figures 1 and 3). The associated cholestatic liver disease phenotypes identified were: ICP (n=5 out of 83 women in the analysis), and gallstone disease (n=10) (Table 3). Some were linked to cholestatic phenotypes previously reported in the literature (n=14), whilst for 19, no phenotype had been previously reported (n=19) (Supplementary Table 4). Likely novel LoF variants were identified (n=3): a frameshift, a splice-acceptor and an introduction of premature stop codon variant. These variants

**Figure 3.**
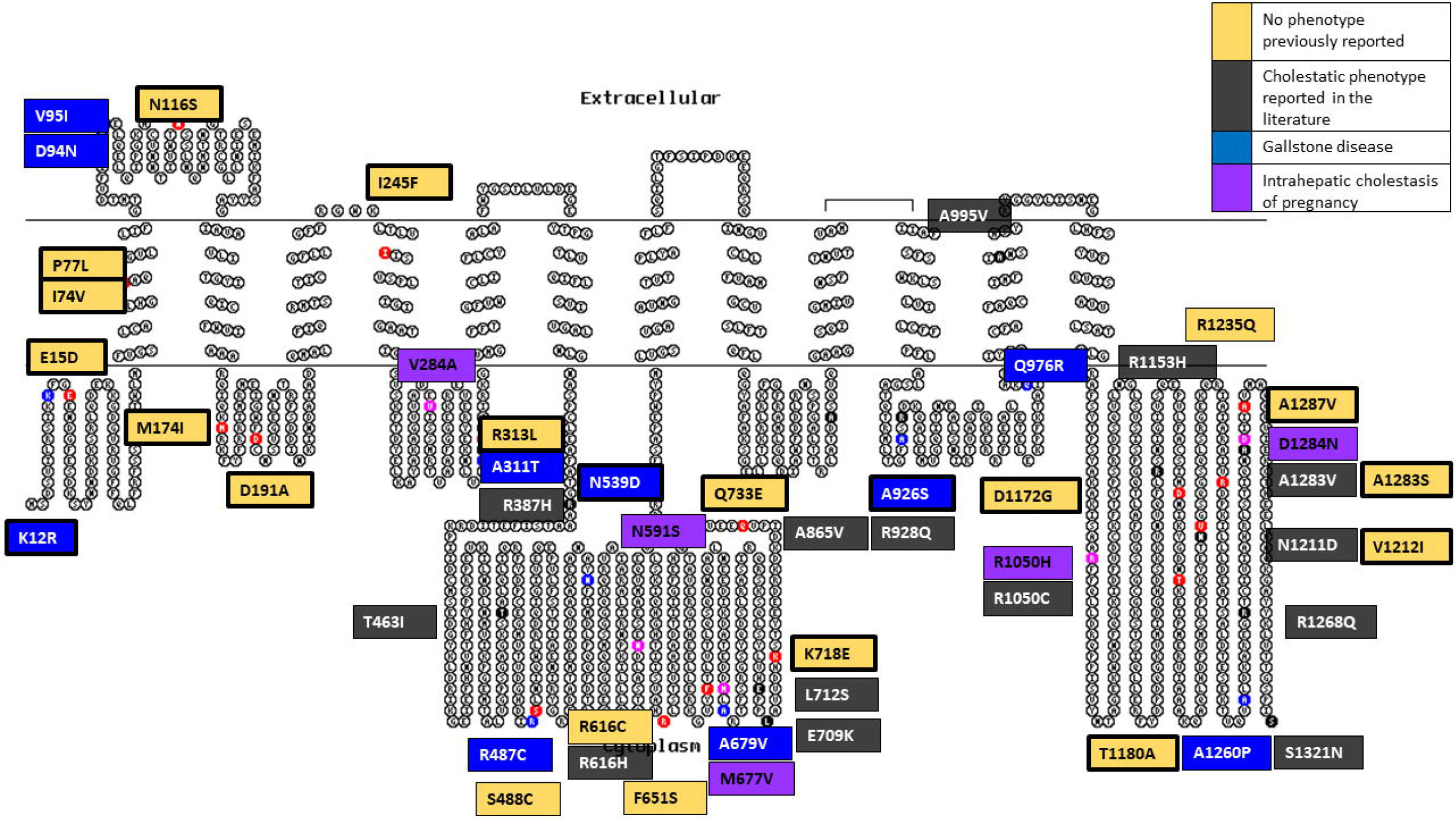
ABCB11 variant summary in a 2-dimensional illustration. Variants are divided into their phenotypic presentation: No phenotype previously reported, cholestatic phenotype reported in the literature, gallstone disease, and intrahepatic cholestasis of pregnancy. Bold border represents variants that are unique to the Genes & Health cohort.

### ATP8B1 variants

We identified a total of 50 *ATP8B1* variants and 31 were included in the final analysis (Figures 1 and 4). There were 7 variants associated with gallstone disease; three appeared to be in linkage disequilibrium (LD) noted in three volunteers: M674T, I577V, and H78Q. The R384H variant was associated with gallstone disease and with an ICP phenotype (in separate individuals, n=2 out of 33 women in the analysis). A further variant was associated with hepatitis-induced liver cirrhosis (I513T), and a final variant (D70N) was associated with liver cirrhosis, secondary malignant neoplasm of liver and bile duct, and gallstone disease (Table 3). In addition, previously reported cholestatic liver disease phenotypes (n=7) and variants with no previous reported phenotype were seen (n=15) (Supplementary Table 5). Finally, we identified 4 novel LoF variants: a frameshift/splice region, frameshift, splice-acceptor/coding-sequence, and stop-gain variant. The latter variant was associated with a gallstone disease phenotype (Table 4).

**Figure 4.**
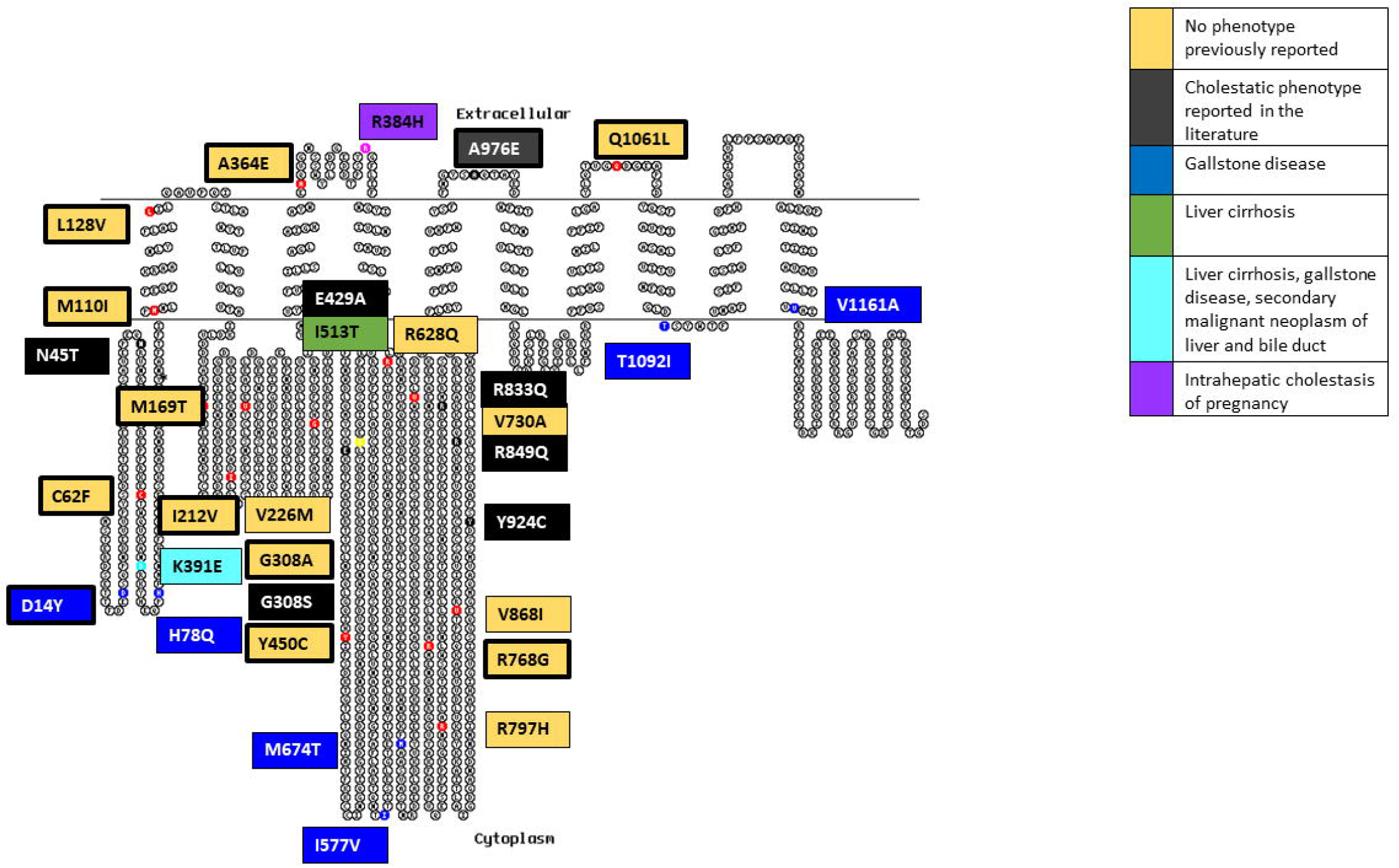
ATP8B1 variant summary in a 2-dimensional illustration. Variants are divided into their phenotypic presentation: No phenotype previously reported, cholestatic phenotype reported in the literature, gallstone disease, liver cirrhosis, Liver cirrhosis, multiple cholestatic phenotype, and intrahepatic cholestasis of pregnancy. Bold border represents variants that are unique to the Genes & Health cohort.

### NR1H4 variants

There were 22 *NR1H4* variants in the Genes & Health cohort and 9 variants in the final analysis (Figure 1 and Supplementary Figure 2). We only identified an ICP phenotype (n=2 out of 33 women in the analysis) (Table 3) and otherwise novel variants that had no previous phenotype reported (n=7) (Supplementary Table 6). Furthermore, one novel LoF variant was identified without demonstrating a phenotype (Table 4).

### TJP2 variants

There were 83 *TJP2* variants identified of which 37 were analysed (Figure 1). People with *TJP2* variants had ICP (n=3 out of 103 women in the analysis), gallstone disease (n=8), previously reported cholestatic liver disease phenotype (n=4), and 22 did not have a previously reported phenotype (Table 3, Supplementary Table 7). There were two novel LoF variants without a clinical phenotype (Table 4).

### Protein structure and molecular modelling

A flow chart illustrating the variants included in this analysis is shown in Supplementary Figure 3. Results of the protein structure and molecular modelling software tools are presented in Supplementary Table 8.

Some novel variants are in regions of transporters for which we can hypothesis a mechanistic impact. Of particular interest are Q1106H in ABCB4 and D191A in ABCB11. These ABC B-family transporters share 48% amino acid identity and are very likely have a common mechanism of action. The two amino acids are conserved in both proteins, and we propose that they are involved in energy transduction through the transporter in order to couple the substrate efflux cycle to the ATP binding and catalysis cycle.

In ABCB4 and ABCB11, two transmembrane domains (TMDs) bind the transport substrates (phosphatidylcholine (PC) and bile acids), respectively. The conformational changes required for substrate transport are driven by ATP hydrolysis at the interface between two nucleotide binding domains (NBDs). The TMDs and NBDs must therefore be intimately coupled, and this is achieved via four ‘coupling helices’ (CH) located at the base of the long intracellular loops extending from the transmembrane alpha helices of the TMDs (Supplementary figure 4A).

Q1106 (ABCB4) and D191 (ABCB11) are particularly interesting because they are located at this interface that is conserved in both ABCB4 and ABCB11. Q1106 is in a groove on the surface of NBD2 where it interacts with CH2 (Supplementary Figure 4B).

In the PC-bound conformation of ABCB4, Q1106 forms a weak electrostatic interaction with the peptide bond of G270. In the ATP-bound conformation (from which PC has most likely been released), Q1106 now interacts with Q272 which illustrates the movement of CH2 and its changing juxtaposition with the NBD during the transport cycle; essentially, a hinge region. The geometry of these interactions will not be preserved if Q1106 is replaced by histidine. In ABCB11, this triad is preserved in Q1150 and G295, with E297 providing a conservative change for the glutamine in CH2 (with respect to formation of an equivalent electrostatic bond with Q1150).

In the sole structural model that we have for ABCB11, D191 is in CH1 where it interacts with Y472 in a surface groove of NBD1 and also, intriguingly, with R946 which is in CH4, suggesting that CH1 and CH4 likely work together in energy transduction through the transporter (Supplementary Figure 4C).

These electrostatic bonds will not be possible if D191 is replaced with alanine. This triad and its bond architecture is perfectly conserved in ABCB4 in the ATP bound conformation through amino acids D166, Y446 and R902. However, in ABCB4 there is also an additional electrostatic interaction between the carbonyl of the D166 peptide bond and the side chain of Q1171. Q1171 (which is conserved in ABCB11) is adjacent to the ABC signature motif ^1172^LSGGQ^1176^ which is involved in coordination of ATP and provides a direct mechanism for how CH1 influences, and responds to, the ATP catalytic cycle of these transporters.

## Discussion

This study identified novel variants implicated in the aetiopathogenesis of cholestatic liver disease that occur uniquely in this British Bangladeshi and Pakistani cohort [39–42]. There have not been any other studies of this magnitude analysing the burden of mutational variation in cholestatic liver disease in a large South Asian cohort. Using a genotype to phenotype approach we discovered novel likely pathogenic variants that appear to be unique to this cohort. We then employed a phenotype to genotype analysis using the ICP phenotype as an exemplar, which offered a pragmatic interrogation of electronic health records to identify rare genetic variants that are likely pathogenic. Thus, this study improves representation of this distinct population especially as prevalence of cholestatic liver disease is increased in the Genes & Health cohort, e.g., 1.54% are affected by ICP compared to white Europeans (0.62%). This study demonstrates the importance of multi-ancestry genomic research and offers the potential of tailored treatment for this population.

In the Genes & Health cohort, out of 194 variants meeting inclusion criteria we identified 53 that had a cholestatic liver disease phenotype reported in their linked EHR. Of those, 16 are unique to this British Bangladeshi and Pakistani population and a large number were predicted to be likely pathogenic or pathogenic based on *in-silico* prediction tools. In addition, there were 35 variants that were previously reported in the literature with a cholestatic phenotype. However, 87 variants had no previously reported phenotype; 67 were novel (34% of all variants analysed in this study) as they were also not previously reported in the GnomAD population database. Despite that, 9 were considered likely pathologic and 5 pathogenic. It is important to consider that heterozygosity as noted in most cases means that they are likely rescued by the wild-type allele but at higher risk of disease in later life or during times of liver stress, e.g. during pregnancy.

Our findings reflect the difficulty with interpretation of rare variants in clinically important genes when there is no previous evidence in the literature or functional data to interpret them further. The ACMG rare variant interpretation guideline[30] provides a standardised analysis pathway. However, it relies in part on the interpretation of the variant in the context of the literature and does not account for specific genes and diseases. It also may not be robust for flexible membrane proteins which do not work by lock and key mechanism. For example, the *ABCB11* variant V444A, considered as benign by the ACMG criteria, has been reported to increase the risk of ICP, hepatitis C disease progression, and drug-induced liver injury although the exact functional mechanisms are not clear yet[43, 44].

By employing computational protein modelling software tools, we were able to identify variants that likely have a significant impact on the conformation of the protein and could therefore be of clinical significance. It is important to bear in mind that all these tools have inherent flaws and are beyond the scope of this paper to discuss in detail. By taking ICP as a cholestatic liver disease example we were able to highlight further difficulties with rare variant interpretation in gestational syndromes as the inherent transient nature of the disease makes variant interpretation challenging. However, ICP is a clinically relevant example as the gestational disease consequences are not just relevant to their current pregnancy but also can result in later hepatobiliary disorders such as cancer, immune-mediated and cardiovascular diseases[45]. In addition, they have a higher gallstone-related morbidity and a strong positive association between ICP and hepatitis C exists as well[46].

## Limitations

The use of electronic health records to determine phenotype is extremely useful but dependent upon appropriate information having been coded. Participants with at-risk variants may not have presented yet with symptoms of disease but still be at high risk of developing complications at a later stage in their life, particularly given that the median age of volunteers in this study was around 45 years. It demonstrates the difficulty with interpreting variants when recalling the genotype first.

## Conclusions

In this study we provide the first comprehensive evaluation of gene candidates associated with cholestatic liver diseases in a unique cohort of British Bangladeshi and Pakistani origin demonstrating the importance of characterising genetic disease in diverse ethnic groups. Our findings have demonstrated the increased mutational burden of cholestatic liver disease in British Bangladeshi and Pakistani people who thus far remain understudied despite their distinct genetic background and increased risk of developing ICP in comparison to other populations. We were able to identify novel variants that have not been previously identified and are likely implicated in disease. We demonstrated the ability to identify participants at risk both by a phenotype or genotype first approach. This demonstrates the importance of providing more personalised care in a clinical setting as identification of high-risk individuals and their family members enables early intervention to prevent adverse outcomes, for example hepato-protective drugs such as UDCA, in addition to hepatic surveillance. Furthermore, it provides the necessary foundation for improved therapy and drug development.

## Data Availability

All data produced in the present study are available upon reasonable request to the authors

## Supplementary Figure legends

**Supplementary Figure 1.**
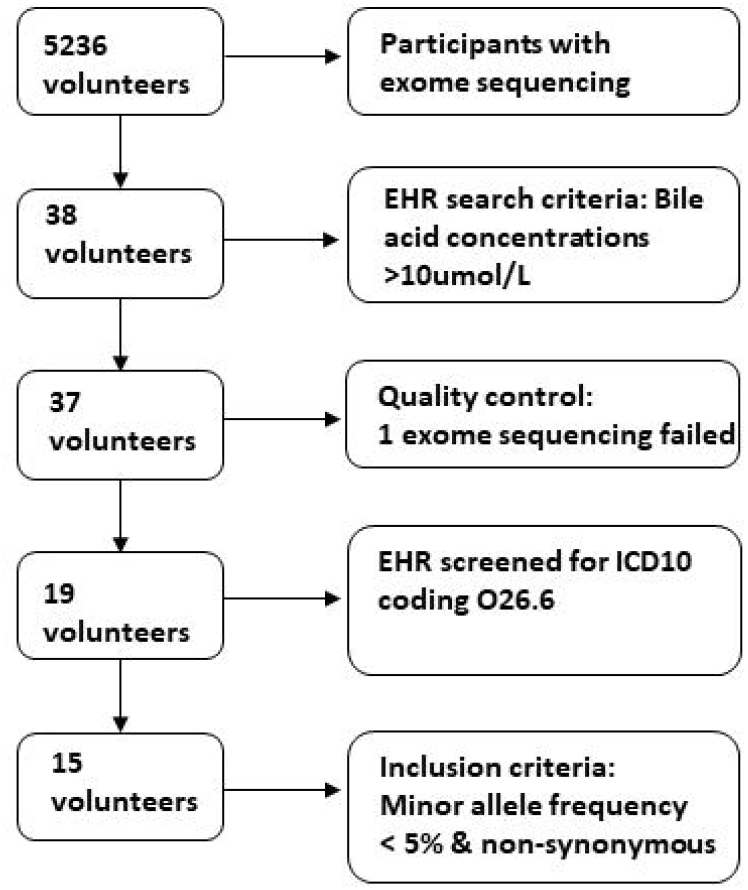
Flow diagram illustrating the patient inclusion for the phenotype to genotype analysis. This diagram illustrates the number of volunteers included with a phenotypic presentation of intrahepatic cholestasis of pregnancy selected from the 5236 exome sequencing data available in the Genes and Health database.

**Supplementary Figure 2.**
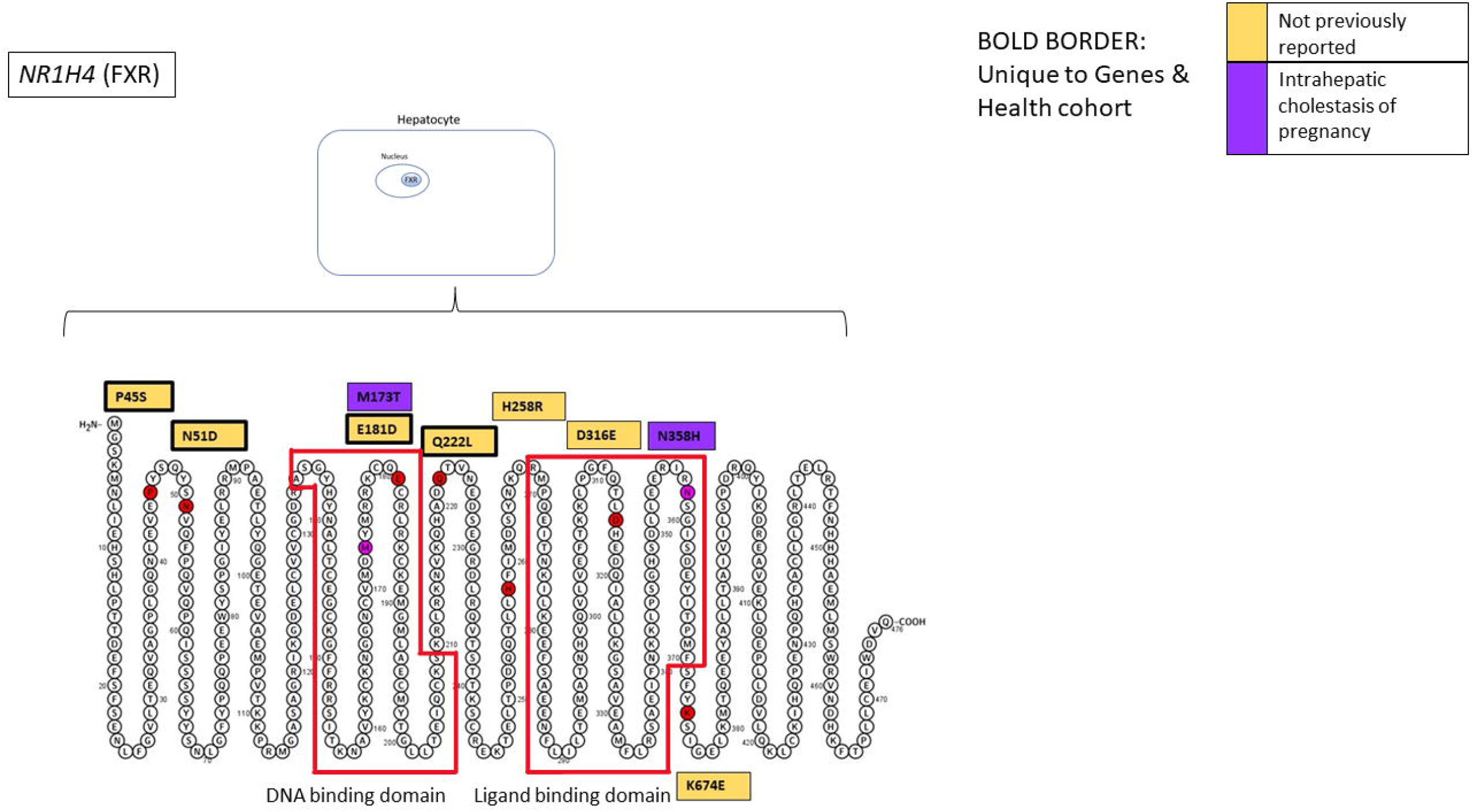
NR1H4 variant summary in a 2-dimensional illustration. NR1H4 also known as FXR is a nuclear receptor within hepatocytes. Variants are divided into their phenotypic presentation: No phenotype previously reported, and intrahepatic cholestasis of pregnancy. Bold border represents variants that are unique to the Genes & Health cohort.

**Supplementary Figure 3.**
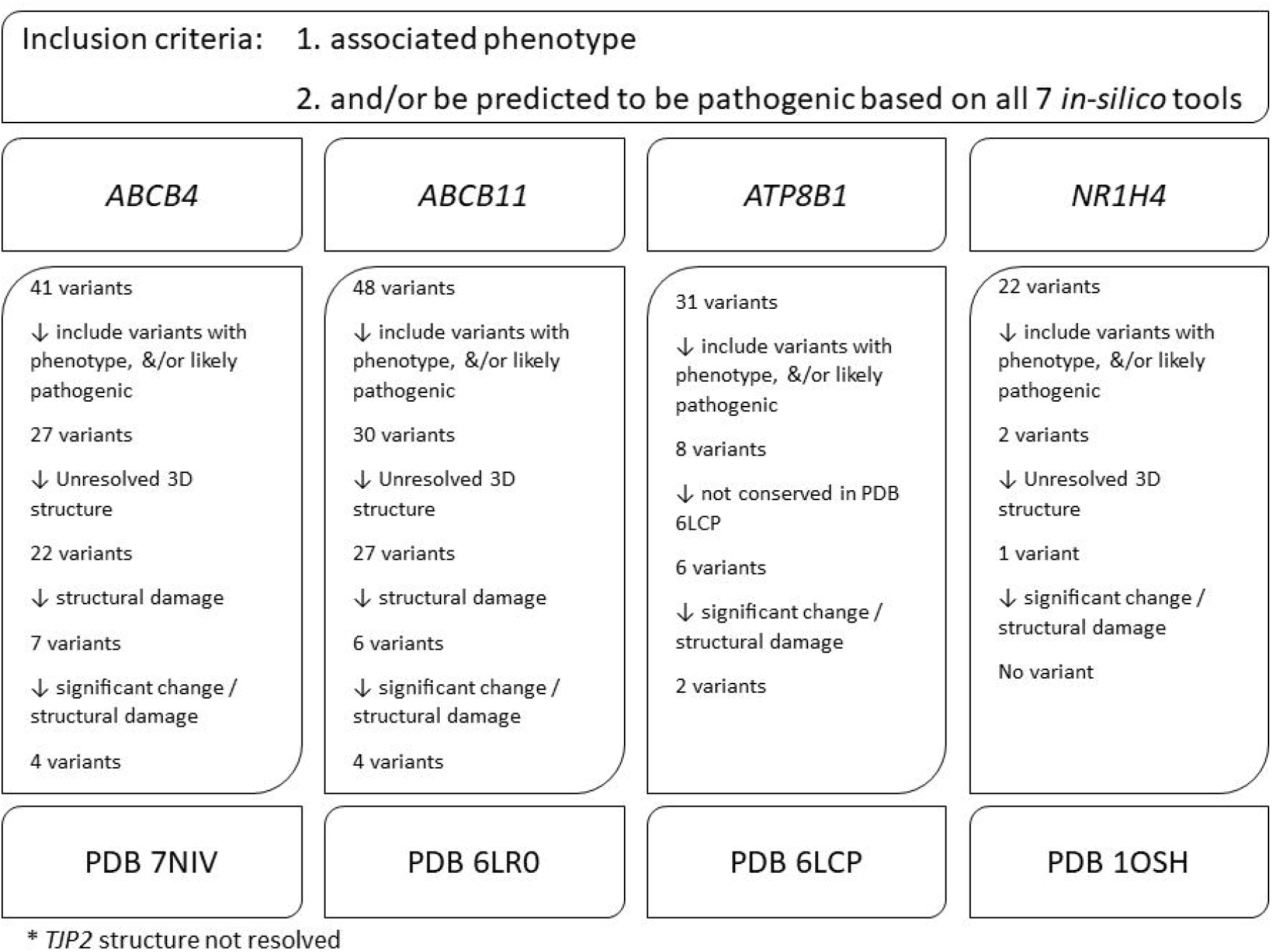
Overall summary of all variants included for the protein modelling in the Genes and Health cohort for all five gene candidates. All variants included for the analysis were modelled and stratified by their predicted structural effect on the 3D structure. *TJP2 structure not included as no 3D structure available.

**Supplementary Figure 4.**
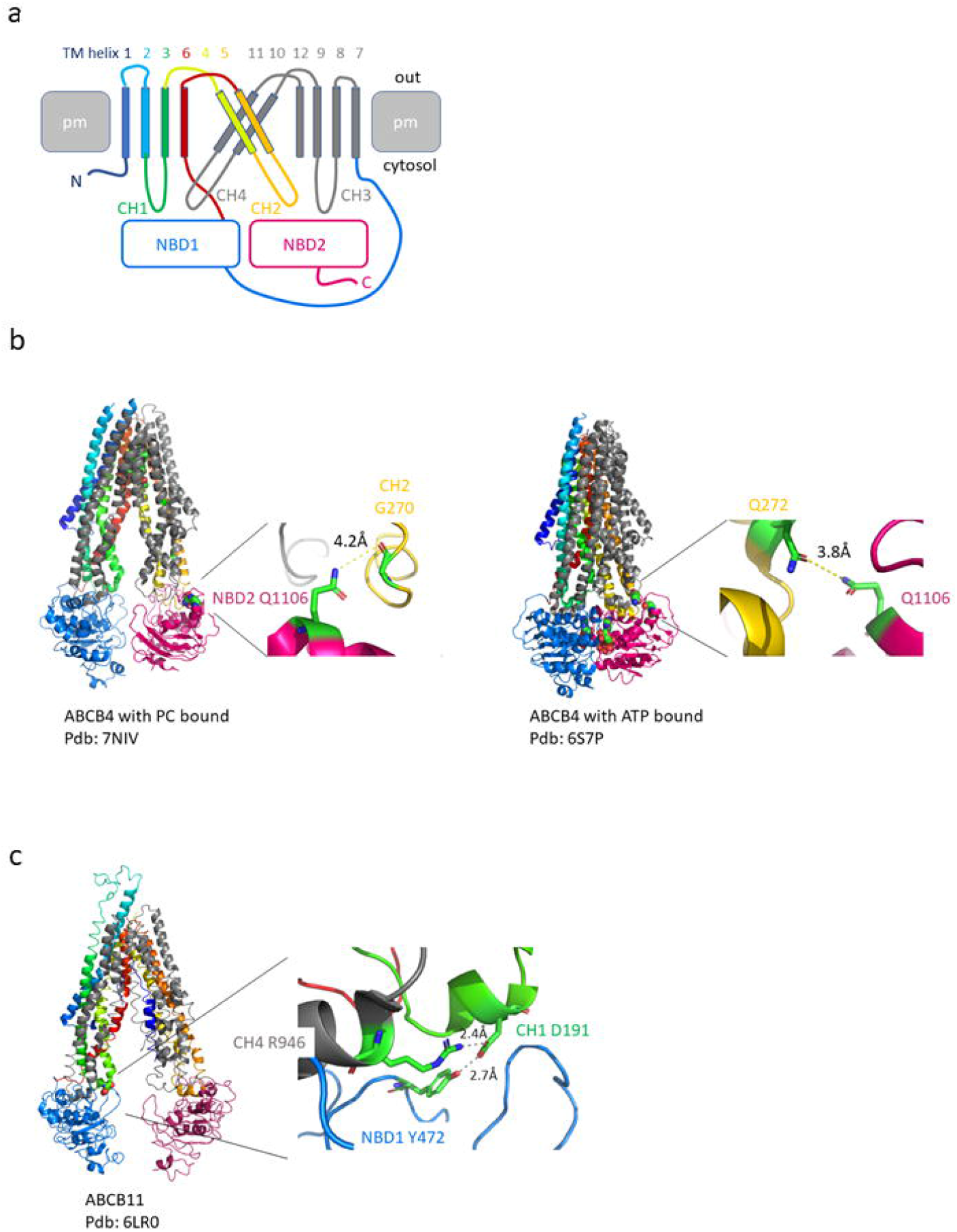
Rare variants at the energy transduction interface between the ligand binding and ATP catalytic domains of ABCB4 and ABCB11. **A. Topological cartoon showing the organisation of the transmembrane domains (TMDs) and the interaction with the nucleotide binding domains (NBDs) of ABCB4 and ABCB11.** Each TMD comprises 6 transmembrane alpha helices (TMH 1-6 of TMD1 in rainbow colouring, TMH 7-12 of TMD2 in grey). The intracellular loops between TMH 2 and 3, 4 and 5, 8 and 9, and 10 and 11 extend into the cytosol to interact with the NBDs (blue and pink squares represent NBD1 and NBD2 respectively). At the base of each of these intracellular loops is a coupling helix (labelled CH1-4) which interdigitate with grooves on the top surface of the NBDs. The coupling helices interact with the surface of the NBDs and are assumed to convey conformational changes in a bidirectional manner: to the NBDs from the TMDs to signal substrate binding to the latter and, vice versa to the TMDs from the NBDs to drive the conformational changes associated with ATP binding (resulting in substrate efflux) and the conformational changes associated with ATP hydrolysis to reset the transporter for binding of the next transport substrate. The domain colour coding is preserved in 4B and 4C. **B. ABCB4 variant Q1106H is in a functionally sensitive location for energy transduction, disrupting the interaction between CH2 and NBD2**. Left panel; cartoon representation of ABCB4 with bound PC. PC is shown in sphere format and coloured elementally (pdb: 7NIV). Q1106 also shown as spheres with elemental colouring is located in a groove on the top surface of NBD2 (pink). The groove is occupied by CH2 formed between TMH4 (yellow) and TMH5 (orange). The distance between the side chain nitrogen (blue) of Q1106 and the nearest atom in the coupling helix, the oxygen (red) of the peptide bond formed by G270 is indicated in the zoomed image where the relevant amino acids are shown in stick format with elemental colouring (nitrogen in blue, oxygen in red, carbon in green). Right panel; cartoon representation of ABCB4 with 2 x Mg^2+^ and 2 x ATP (pdb: 6S7P). Q1106 is now in close proximity to Q272 in CH2. The distance between the side chain nitrogen (blue) of Q1106 and the oxygen (red) of the Q272 is indicated in the zoomed image. Interatom distances were measured in Pymol. **C. ABCB11 variant D191A is in a functionally sensitive location for energy transduction, disrupting the interaction between CH1 and NBD1**. Left panel; cartoon representation of ABCB11 (pdb: 6LR0). D191 which is shown as spheres with elemental colouring is located in CH1 where it interacts with both Y472 on the top surface of NBD1 (blue) and R946 from CH4. Minimum distances between the oxygens of the carboxylic acid side chain of D191 and the oxygen of Y472 and the nitrogen of the guanidinium group of R946 are shown in the zoomed image where the relevant amino acids are shown in stick format with elemental colouring as above.

