## Supplemental tables for "Rare variant contribution to cholestatic liver disease in a South Asian population in the United Kingdom"

**Supplementary table 1. *Summary of all variants identified including clinical, literature review, protein function and in-silico variant prediction.***

|  | **SNV** | | | | | **Protein change** | | **Protein function prediction** | | | ***In-silico* variant prediction** | | | | | | | |
| --- | --- | --- | --- | --- | --- | --- | --- | --- | --- | --- | --- | --- | --- | --- | --- | --- | --- | --- |
|  | **CHR** | **POS** | **REF** | **ALT** |  | | **Conserved** | | **Disease propensity** | **Other variants in region** | **Varsome** | **SIFT** | **PolyPhen** | **CADD** | **Revel** | **MetaLR** | **Meta_SVM** | **M_CAP** |
| ABCB4 | chr7 | 87402176 | C | T | G1254S | | 1 | | 1 | 0 | 1 | deleterious(0) | probably_damaging(1) | 26.3 | 0.94802 | D | D | D |
|  | chr7 | 87408168 | G | A | P1050S | | 1 | | 0 | 1 | 1 | tolerated(0.21) | possibly_damaging(0.767) | 22.8 | 0.6331 | D | D | D |
|  | chr7 | 87417497 | C | T | A833T | | 0 | | 0 | 1 | 1 | deleterious(0) | probably_damaging(1) | 27 | 0.88496 | D | D | D |
|  | chr7 | 87440230 | T | C | N510S | | 1 | | 0 | 1 | 1 | deleterious(0.02) | possibly_damaging(0.854) | 23.1 | 0.82556 | D | D | D |
|  | chr7 | 87426810 | C | G | Q668H | | 0 | | 0 | 0 | 3 | tolerated(0.31) | benign(0) | 6.848 | 0.60481 | T | T | D |
|  | chr7 | 87406364 | C | T | R1137Q | | 0 | | 1 | 0 | 1 | deleterious(0.02) | possibly_damaging(0.46) | 26.2 | 0.87877 | D | D | D |
|  | chr7 | 87420029 | C | T,A | R788L | | 0 | | 1 | 1 | 1 | deleterious(0) | probably_damaging(0.982) | 23.6 | 0.88684 | T | T | D |
|  | chr7 | 87426862 | G | T | T651N | | 0 | | 0 | 1 | 3 | tolerated(0.43) | benign(0.038) | 0.999 | 0.423 | T | T | D |
|  | chr7 | 87443722 | T | C | K391E | | 1 | | 0 | 1 | 3 | tolerated(0.23) | benign(0.35) | 19.78 | 0.66922 | D | T | D |
|  | chr7 | 87418539 | C | T | G826R | | 1 | | 1 | 0 | 1 | deleterious(0) | probably_damaging(1) | 33 | 0.95499 | D | D | D |
|  | chr7 | 87426786 | C | T | M676I | | 0 | | 0 | 0 | 2 | tolerated(0.44) | benign(0) | 1.525 | 0.51995 | T | T | D |
|  | chr7 | 87426758 | C | T | D686N | | 0 | | 0 | 1 | 2 | tolerated(0.22) | benign(0) | 1.98 | 0.32697 | T | T | D |
|  | chr7 | 87454542 | T | C | M113V | | 1 | | 0 | 1 | 1 | deleterious(0.04) | benign(0.079) | 22.8 | 0.80713 | T | T | D |
|  | chr7 | 87420068 | G | A | T775M | | 1 | | 1 | 1 | 2 | tolerated(0.5) | probably_damaging(0.944) | 22.7 | 0.8124 | D | T | D |
|  | chr7 | 87439833 | A | G | F522S | | 1 | | 1 | 1 | 1 | deleterious(0) | probably_damaging(0.92) | 28.3 | 0.96627 | D | D | D |
|  | chr7 | 87440360 | A | C | Y467D | | 1 | | 1 | 1 | 1 | deleterious(0.01) | possibly_damaging(0.674) | 23.8 | 0.83394 | D | D | D |
|  | chr7 | 87449993 | C | G | G270R | | 1 | | 1 | 0 | 1 | deleterious(0.01) | probably_damaging(0.933) | 26.9 | 0.92964 | D | D | D |
|  | chr7 | 87426778 | T | C | K679R | | 0 | | 0 | 0 | 2 | tolerated(0.6) | benign(0) | 4.349 | 0.45696 | T | T | D |
|  | chr7 | 87406456 | C | G | Q1106H | | 2 | | 0 | 1 | 1 | tolerated(0.09) | benign(0.085) | 22.5 | 0.62767 | D | D | D |
|  | chr7 | 87431528 | C | T | R590Q | | 1 | | 1 | 1 | 4 | deleterious(0.01) | probably_damaging(0.967) | 31 | 0.89829 | D | D | . |
|  | chr7 | 87453110 | C | A | G124C | | 1 | | 1 | 1 | 1 | deleterious(0) | probably_damaging(0.998) | 29.4 | 0.98549 | D | D | D |
|  | chr7 | 87462827 | G | C | L73V | | 1 | | 0 | 1 | 1 | tolerated(0.3) | benign(0.29) | 14.12 | 0.60123 | D | T | D |
|  | chr7 | 87426841 | C | A | W658L | | 0 | | 1 | 1 | 2 | tolerated(0.65) | benign(0) | 14.87 | 0.5927 | T | T | D |
|  | chr7 | 87406437 | C | T | G1113R | | 1 | | 1 | 1 | 1 | deleterious(0) | probably_damaging(1) | 31 | 0.97885 | D | D | D |
|  | chr7 | 87431565 | C | A | A578S | | 1 | | 0 | 1 | 1 | deleterious(0) | probably_damaging(0.936) | 33 | 0.88963 | D | D | D |
|  | chr7 | 87406316 | A | T | I1153K | | 1 | | 1 | 0 | 1 | deleterious(0) | possibly_damaging(0.887) | 27 | 0.99443 | D | D | D |
|  | chr7 | 87406404 | A | G | C1124R | | 1 | | 1 | 0 | 1 | tolerated(0.1) | probably_damaging(0.99) | 25.9 | 0.93475 | D | D | D |
|  | chr7 | 87408039 | C | T | V1093M | | 0 | | 0 | 1 | 1 | tolerated(0.05) | probably_damaging(0.962) | 26.1 | 0.9211 | D | D | D |
|  | chr7 | 87409306 | G | T | A1004E | | 0 | | 1 | 0 | 2 | deleterious(0) | probably_damaging(0.971) | 28.4 | 0.96919 | D | D | D |
|  | chr7 | 87451757 | C | A,T | V192I | | 0 (0) | | 1 (0) | 1 (1) | 1 | tolerated(1) | benign(0.005) | 16.12 | 0.52596 | T | T | D |
|  | chr7 | 87402223 | T | C | N1238S | | 1 | | 0 | 0 | 2 | deleterious(0.04) | possibly_damaging(0.559) | 15.09 | 0.60717 | T | T | D |
|  | chr7 | 87422179 | A | G | I753T | | 0 | | 0 | 1 | 2 | tolerated(0.23) | benign(0.003) | 19.75 | 0.59021 | T | T | D |
|  | chr7 | 87422195 | G | C | Q748E | | 0 | | 0 | 1 | 2 | tolerated(0.19) | benign(0.02) | 21.2 | 0.63418 | T | D | D |
|  | chr7 | 87472653 | T | C | K35E | | 0 | | 0 | 1 | 2 | tolerated(0.94) | benign(0.003) | 15.86 | 0.49989 | T | T | D |
|  | chr7 | 87408137 | T | C | K1060R | | 0 | | 0 | 1 | 2 | tolerated(0.12) | benign(0.062) | 22.7 | 0.48707 | D | T | D |
|  | chr7 | 87426763 | T | A | E684V | | 0 | | 1 | 0 | 2 | tolerated(0.06) | benign(0.007) | 16.25 | 0.53625 | D | T | D |
|  | chr7 | 87444956 | A | G | I342T | | 1 | | 0 | 1 | 3 | deleterious(0.02) | benign(0.061) | 23.4 | 0.74093 | T | T | D |
|  | chr7 | 87453040 | C | T | R147K | | 0 | | 0 | 1 | 2 | tolerated(1) | benign(0) | 9.241 | 0.5602 | T | T | D |
|  | chr7 | 87402265 | C | T | R1224H | | 1 | | 1 | 0 | 1 | deleterious(0.01) | probably_damaging(0.994) | 31 | 0.9328 | D | D | D |
|  | chr7 | 87418605 | C | T | D804N | | 1 | | 0 | 0 | 1 | deleterious(0.01) | possibly_damaging(0.767) | 32 | 0.92438 | D | D | D |
|  | chr7 | 87452957 | T | C | T175A | | 1 | | 0 | 1 | 3 | tolerated(0.05) | possibly_damaging(0.615) | 24 | 0.83339 | T | D | . |
| ABCB11 | chr2 | 168923738 | C | T | D1284N | | 0 | | 0 | 1 | 1 | deleterious(0.03) | benign(0.057) | 23.5 | 0.76289 | T | T | D |
|  | chr2 | 168932441 | C | T | R1050H | | 0 | | 1 | 1 | 2 | tolerated(0.9) | probably_damaging(0.912) | 23.2 | 0.79395 | D | D | D |
|  | chr2 | 168990858 | A | G | V284A | | 2 | | 0 | 1 | 1 | tolerated(0.15) | probably_damaging(0.93) | 24.6 | 0.8887 | D | D | D |
|  | chr2 | 168970082 | T | C | N591S | | 1 | | 0 | 1 | 4 | tolerated(0.06) | possibly_damaging(0.883) | 24 | 0.61416 | T | T | . |
|  | Hom (n) | 99 | Het (n) | 760 |  |  |  |  |  |  |  |  |  |  |  |  |  |  |
|  | chr2 | 168968473 | T | C | M677V | | 0 | | 0 | 1 | 4 | tolerated(0.46) | benign(0) | 0.716 | 0.50616 | T | T | . |
|  | Het (n) | 99 | Hom (n) | 3 |  |  |  |  |  |  |  |  |  |  |  |  |  |  |
|  | chr2 | 168923810 | C | G | A1260P | | 1 | | 1 | 1 | 1 | deleterious(0) | probably_damaging(0.949) | 27.3 | 0.97698 | D | D | D |
|  | chr2 | 168935313 | T | C | Q976R | | 1 | | 0 | 1 | 2 | tolerated(0.58) | benign(0.001) | 0.622 | 0.42115 | T | T | D |
|  | chr2 | 168936268 | C | A | A926S | | 0 | | 0 | 1 | 1 | tolerated(0.06) | probably_damaging(0.976) | 23.6 | 0.81356 | D | D | D |
|  | chr2 | 168968466 | G | A | A679V | | 0 | | 0 | 0 | 2 | tolerated(0.29) | benign(0.001) | 4.859 | 0.59146 | T | T | D |
|  | chr2 | 168971870 | T | C | N539D | | 1 | | 0 | 1 | 2 | tolerated(1) | benign(0.034) | 19.33 | 0.62325 | T | T | D |
|  | chr2 | 168972026 | G | A | R487C | | 1 | | 1 | 1 | 1 | deleterious(0) | probably_damaging(0.991) | 33 | 0.93631 | D | D | D |
|  | chr2 | 168986262 | C | T | A311T | | 0 | | 0 | 0 | 1 | deleterious(0) | probably_damaging(0.975) | 26 | 0.90885 | D | D | D |
|  | chr2 | 169013378 | C | T | V95I | | 0 | | 0 | 1 | 3 | tolerated(1) | benign(0.003) | 0.919 | 0.43033 | T | T | D |
|  | chr2 | 169013381 | C | T | D94N | | 0 | | 0 | 1 | 1 | tolerated(0.13) | possibly_damaging(0.815) | 22.7 | 0.72016 | T | T | D |
|  | chr2 | 169018091 | T | C | K12R | | 0 | | 0 | 0 | 2 | tolerated_low_confidence(0.19) | benign(0.007) | 20.4 | 0.47225 | T | T | D |
|  | chr2 | 168923741 | C | A | A1283S | | 1 | | 0 | 1 | 2 | tolerated(0.3) | benign(0.005) | 18.14 | 0.70014 | D | D | D |
|  | chr2 | 168923785 | C | T | R1268Q | | 1 | | 1 | 0 | 1 | deleterious(0) | probably_damaging(0.999) | 29.4 | 0.97997 | D | D | D |
|  | chr2 | 168924718 | C | T | R1235Q | | 1 | | 1 | 1 | 1 | deleterious(0.02) | probably_damaging(1) | 28.9 | 0.86753 | D | D | D |
|  | chr2 | 168927259 | T | C | D1172G | | 1 | | 0 | 0 | 1 | deleterious(0.01) | probably_damaging(0.957) | 29.7 | 0.99039 | D | D | D |
|  | chr2 | 168927316 | C | T | R1153H | | 2 | | 1 | 1 | 1 | deleterious(0.01) | probably_damaging(1) | 27.5 | 0.97512 | D | D | D |
|  | chr2 | 168932442 | G | A | R1050C | | 1 | | 1 | 1 | 1 | deleterious(0.02) | probably_damaging(0.939) | 32 | 0.97475 | D | D | D |
|  | chr2 | 168935256 | G | A | A995V | | 0 | | 0 | 1 | 1 | tolerated(0.1) | possibly_damaging(0.833) | 23.7 | 0.78535 | D | D | D |
|  | chr2 | 168936261 | C | T | R928Q | | 1 | | 1 | 1 | 2 | tolerated(0.55) | benign(0.003) | 13.29 | 0.80058 | T | T | D |
|  | chr2 | 168964232 | T | C | K718E | | 0 | | 0 | 1 | 2 | tolerated(0.3) | benign(0) | 15.19 | 0.60123 | T | T | D |
|  | chr2 | 168969514 | C | T | R616H | | 1 | | 1 | 0 | 1 | deleterious(0) | probably_damaging(0.999) | 24.9 | 0.91611 | D | D | D |
|  | chr2 | 168972022 | G | C | S488C | | 1 | | 0 | 0 | 1 | deleterious(0) | probably_damaging(0.935) | 26 | 0.8124 | D | D | D |
|  | chr2 | 168973761 | G | A | T463I | | 0 | | 0 | 1 | 1 | deleterious(0) | probably_damaging(0.991) | 28.3 | 0.98834 | D | D | D |
|  | chr2 | 168995438 | C | T | M174I | | 1 | | 0 | 0 | 2 | tolerated(1) | benign(0) | 8.761 | 0.5046 | T | T | D |
|  | chr2 | 169013314 | T | C | N116S | | 0 | | 0 | 1 | 1 | tolerated(0.07) | probably_damaging(0.965) | 22.9 | 0.81761 | D | D | D |
|  | chr2 | 169013431 | G | A | P77L | | 0 | | 0 | 1 | 1 | deleterious(0) | probably_damaging(0.976) | 27.5 | 0.95935 | D | D | D |
|  | chr2 | 168923626 | C | T | S1321N | | 1 | | 0 | 0 | 2 | deleterious_low_confidence(0) | probably_damaging(0.998) | 24.7 | 0.81761 | D | D | D |
|  | chr2 | 168923728 | G | A | A1287V | | 1 | | 0 | 0 | 2 | tolerated(0.58) | benign(0.127) | 20.5 | 0.52296 | T | T | D |
|  | chr2 | 168923740 | G | A | A1283V | | 1 | | 0 | 1 | 1 | deleterious(0) | benign(0.311) | 24.7 | 0.84535 | D | D | D |
|  | chr2 | 168924788 | C | T | V1212I | | 1 | | 0 | 0 | 1 | deleterious(0.01) | probably_damaging(0.999) | 26 | 0.80238 | D | D | D |
|  | chr2 | 168924791 | T | C | N1211D | | 1 | | 0 | 0 | 2 | tolerated(0.28) | benign(0.052) | 22.3 | 0.58135 | T | T | D |
|  | chr2 | 168927236 | T | C | T1180A | | 2 | | 0 | 0 | 2 | deleterious(0.01) | benign(0.338) | 23.3 | 0.64157 | T | T | D |
|  | chr2 | 168944621 | G | A | A865V | | 2 | | 0 | 1 | 4 | tolerated(0.07) | possibly_damaging(0.647) | 26 | 0.81818 | D | D | . |
|  | chr2 | 168958110 | G | C | Q733E | | 1 | | 0 | 0 | 2 | tolerated(1) | benign(0) | 8.506 | 0.40989 | T | T | D |
|  | chr2 | 168964249 | A | G | L712S | | 0 | | 1 | 0 | 2 | tolerated(0.42) | benign(0.001) | 16.74 | 0.5602 | T | T | D |
|  | chr2 | 168964259 | C | T | E709K | | 0 | | 1 | 0 | 2 | tolerated(0.24) | benign(0.036) | 22.2 | 0.54201 | T | T | D |
|  | chr2 | 168969409 | A | G | F651S | | 0 | | 1 | 0 | 1 | tolerated(0.08) | probably_damaging(0.999) | 25.8 | 0.83339 | T | T | D |
|  | chr2 | 168969515 | G | A | R616C | | 1 | | 1 | 1 | 1 | deleterious(0) | probably_damaging(1) | 28.9 | 0.97327 | D | D | D |
|  | chr2 | 168979903 | C | T | R387H | | 0 | | 1 | 0 | 2 | deleterious(0) | possibly_damaging(0.661) | 26.2 | 0.89694 | D | D | D |
|  | chr2 | 168986255 | C | A | R313L | | 0 | | 1 | 0 | 2 | deleterious(0.05) | benign(0.007) | 22.3 | 0.54769 | T | T | D |
|  | chr2 | 168993761 | T | A | I245F | | 0 | | 1 | 1 | 1 | deleterious(0) | possibly_damaging(0.784) | 26.7 | 0.88823 | D | D | D |
|  | chr2 | 168995388 | T | G | D191A | | 0 | | 1 | 1 | 1 | deleterious(0) | probably_damaging(1) | 28.3 | 0.98183 | D | D | D |
|  | chr2 | 169013441 | T | C | I74V | | 1 | | 0 | 0 | 2 | tolerated(0.35) | benign(0.001) | 2.838 | 0.55468 | T | T | D |
|  | chr2 | 169018081 | C | G | E15D | | 0 | | 0 | 0 | 2 | tolerated(0.08) | benign(0.318) | 13.66 | 0.64157 | T | T | D |
| ATP8B1 | chr18 | 57691876 | C | T | R384H | | 2 | | 1 | 1 | 2 | deleterious(0.03) | possibly_damaging(0.714) | 24.4 | 0.7727 | D | D | D |
|  | chr18 | 57650416 | A | G | V1161A | | 0 | | 0 | . | 2 | deleterious(0.04) | benign(0.051) | 24.2 | 0.51233 | T | T | D |
|  | chr18 | 57652159 | G | A | T1092I | | 0 | | 0 | 0 | 2 | tolerated(0.08) | probably_damaging(0.977) | 25.9 | 0.82103 | T | T | D |
|  | chr18 | 57731768 | C | A | D14Y | | 1 | | 1 | . | 2 | deleterious_low_confidence(0) | possibly_damaging(0.641) | 24.9 | 0.35136 | T | T | T |
|  | chr18 | 57684128 | A | G | I513T | | 0 | | 0 | 0 | 2 | deleterious(0.02) | possibly_damaging(0.6) | 24.3 | 0.66064 | T | T | D |
|  | chr18 | 57655198 | G | T | A976E | | 1 | | 1 | 0 | 2 | deleterious(0) | possibly_damaging(0.753) | 28.2 | 0.83725 | T | T | D |
|  | chr18 | 57661279 | C | T | V868I | | 1 | | 0 | 1 | 2 | deleterious(0) | probably_damaging(0.996) | 26.6 | 0.83394 | D | D | D |
|  | chr18 | 57661335 | C | T | R849Q | | 0 | | 1 | . | 2 | tolerated(0.4) | benign(0.003) | 2.507 | 0.39643 | T | T | D |
|  | chr18 | 57662599 | T | C | R768G | | 0 | | 0 | 1 | 2 | tolerated(0.08) | benign(0.325) | 23.3 | 0.54344 | T | T | T |
|  | chr18 | 57668449 | A | G | V730A | | 1 | | 0 | 1 | 2 | deleterious(0) | benign(0.243) | 24.6 | 0.93901 | D | D | D |
|  | chr18 | 57688379 | T | C | Y450C | | 1 | | 1 | 1 | 2 | deleterious(0) | probably_damaging(0.998) | 31 | 0.93475 | D | D | D |
|  | chr18 | 57691936 | G | T | A364E | | 0 | | 1 | 0 | 2 | tolerated(0.91) | benign(0.021) | 19.68 | 0.38257 | T | T | T |
|  | chr18 | 57695188 | C | G | G308A | | 2 | | 0 | 1 | 2 | deleterious(0) | probably_damaging(1) | 25 | 0.95572 | D | D | D |
|  | chr18 | 57695189 | C | T | G308S | | 2 | | 1 | 1 | 2 | deleterious(0) | probably_damaging(1) | 26.4 | 0.9833 | D | D | D |
|  | chr18 | 57697640 | C | T | V226M | | 1 | | 0 | 1 | 2 | deleterious(0) | probably_damaging(0.999) | 25.2 | 0.9579 | D | D | D |
|  | chr18 | 57697682 | T | C | I212V | | 1 | | 0 | 1 | 2 | tolerated(0.29) | benign(0.038) | 17.71 | 0.50616 | T | T | T |
|  | chr18 | 57701087 | A | G | M169T | | 1 | | 0 | 0 | 2 | tolerated(0.08) | benign(0.119) | 22.9 | 0.70994 | T | T | D |
|  | chr18 | 57704618 | C | G | M110I | | 1 | | 0 | 0 | 2 | tolerated(0.26) | benign(0) | 20.9 | 0.63842 | T | T | T |
|  | chr18 | 57706584 | C | A | C62F | | 0 | | 1 | . | 2 | tolerated(0.63) | benign(0.029) | 17.92 | 0.66639 | T | T | D |
|  | chr18 | 57652563 | T | A | Q1061L | | 0 | | 0 | 0 | 2 | tolerated(0.2) | benign(0.261) | 23.3 | 0.45868 | T | T | T |
|  | chr18 | 57655354 | T | C | Y924C | | 0 | | 1 | 1 | 2 | deleterious(0) | possibly_damaging(0.603) | 25.6 | 0.94015 | D | D | D |
|  | chr18 | 57662511 | C | T | R797H | | 0 | | 1 | 1 | 2 | tolerated(0.52) | benign(0.017) | 22.4 | 0.41556 | T | T | T |
|  | chr18 | 57671517 | C | T | R628Q | | 1 | | 1 | 1 | 1 | deleterious(0.01) | probably_damaging(0.998) | 31 | 0.87539 | D | D | D |
|  | chr18 | 57688442 | T | G | E429A | | 0 | | 0 | 0 | 3 | tolerated(0.42) | benign(0.003) | 22.4 | 0.36411 | T | T | T |
|  | chr18 | 57704566 | G | C | L128V | | 2 | | 0 | 1 | 2 | tolerated(0.06) | probably_damaging(0.991) | 23.8 | 0.74235 | T | T | D |
|  | chr18 | 57731674 | T | G | N45T | | 0 | | 0 | . | 4 | tolerated_low_confidence(0.11) | benign(0.112) | 15.45 | 0.28106 | T | T | T |
|  | chr18 | 57669394 | A | G | M674T | | 0 | | 0 | 1 | 4 | tolerated(0.5) | benign(0) | 12.04 | 0.42115 | T | T | . |
|  | Het (n) | 62 | Hom (n) | 2 |  |  |  |  |  |  |  |  |  |  |  |  |  |  |
|  | chr18 | 57706535 | G | C | H78Q | | 0 | | 0 | . | 4 | tolerated(0.66) | benign(0) | 0.002 | 0.16033 | T | T | . |
|  | Het (n) | 47 | Hom (n) | 1 |  |  |  |  |  |  |  |  |  |  |  |  |  |  |
|  | chr18 | 57674924 | T | C | I577V | | 1 | | 0 | 1 | 4 | tolerated(0.11) | possibly_damaging(0.787) | 20.3 | 0.57094 | T | T | . |
|  | chr18 | 57706561 | C | T | D70N | | 1 | | 0 | . | 2 | tolerated(0.23) | probably_damaging(0.926) | 23.7 | 0.52745 | T | T | D |
|  | Het (n) | 24 | Hom (n) | 1 |  |  |  |  |  |  |  |  |  |  |  |  |  |  |
|  | chr18 | 57661383 | C | T | R833Q | | 0 | | 1 | 1 | 4 | tolerated(0.55) | benign(0) | 16.99 | 0.23497 | T | T | . |
| NR1H4 | chr12 | 100532530 | T | C | M173T | | 1 | | 0 | 1 | 4 | deleterious(0) | probably_damaging(0.989) | 25.6 | 0.95133 | D | D | D |
|  | chr12 | 100540812 | A | C | N358H | | 2 | | 0 | 0 | 2 | tolerated(0.31) | benign(0.417) | 22.6 | 0.67661 | D | D | D |
|  | chr12 | 100510849 | A | G | N51D | | 0 | | 0 | . | 2 | deleterious(0.02) | benign(0.015) | 20.1 | 0.5491 | D | T | D |
|  | chr12 | 100536552 | A | G | H258R | | 0 | | 0 | 0 | 2 | tolerated(0.16) | benign(0) | 14.75 | 0.4655 | D | T | D |
|  | chr12 | 100540688 | C | G | D316E | | 0 | | 0 | 0 | 2 | deleterious(0.05) | probably_damaging(0.989) | 23.2 | 0.86453 | D | D | D |
|  | chr12 | 100561926 | A | G | K374E | | 1 | | 0 | 0 | 2 | deleterious(0.01) | probably_damaging(0.997) | 27.6 | 0.92804 | D | D | D |
|  | chr12 | 100510831 | C | T | P45S | | 1 | | 0 | . | 2 | tolerated(0.11) | benign(0) | 7.036 | 0.44648 | D | T | D |
|  | chr12 | 100532555 | G | T | E181D | | 2 | | 0 | 1 | 2 | tolerated(0.3) | benign(0.024) | 16.04 | 0.70014 | D | T | D |
|  | chr12 | 100534956 | A | T | Q222L | | 0 | | 0 | 0 | 2 | tolerated(0.67) | benign(0.015) | 19.92 | 0.66064 | D | T | D |
| TJP2 | chr9 | 69236102 | A | G | T377A | | 1 | | 0 | 0 | 2 | tolerated(0.44) | benign(0.167) | 19.82 | 0.12947 | T | T | T |
|  | chr9 | 69205232 | G | A | R21H | | 0 | | 1 | 0 | 4 | tolerated(0.55) | benign(0.003) | 1.869 | 0.06539 | T | T | . |
|  | Het (n) | 443 | Hom (n) | 16 |  |  |  |  |  |  |  |  |  |  |  |  |  |  |
|  | chr9 | 69220926 | C | A | Q105K | | 0 | | 0 | 1 | 4 | tolerated(1) | benign(0.087) | 6.897 | 0.19598 | T | T | . |
|  | chr9 | 69212579 | A | G | Q8R | | 1 | | 0 | 0 | 2 | deleterious(0) | probably_damaging(0.992) | 26.2 | 0.83172 | T | T | D |
|  | chr9 | 69218289 | C | A | T68N | | 0 | | 0 | 0 | 2 | deleterious(0.02) | possibly_damaging(0.765) | 24.2 | 0.48053 | T | T | T |
|  | chr9 | 69221068 | C | T | P152L | | 0 | | 0 | . | 3 | tolerated(0.5) | benign(0.007) | 13.16 | 0.0584 | T | T | T |
|  | chr9 | 69227812 | C | T | R178C | | 0 | | 1 | 0 | 2 | deleterious(0) | probably_damaging(0.91) | 26.7 | 0.37035 | T | T | D |
|  | chr9 | 69229220 | G | A | R255H | | 0 | | 1 | . | 2 | tolerated(0.07) | benign(0.029) | 23.7 | 0.04481 | T | T | T |
|  | chr9 | 69237065 | G | C | R461P | | 1 | | 1 | 0 | 2 | deleterious(0) | probably_damaging(1) | 31 | 0.75074 | T | T | D |
|  | chr9 | 69248118 | C | T | T902M | | 2 | | 1 | . | 2 | deleterious(0) | probably_damaging(1) | 26.1 | 0.6757 | T | T | D |
|  | chr9 | 69251321 | G | A | R1070K | | 0 | | 0 | . | 2 | tolerated_low_confidence(0.11) | benign(0.019) | 27.6 | 0.5046 | T | T | D |
|  | chr9 | 69174379 | G | C | V3L | | 1 | | 0 | 0 | 2 | deleterious_low_confidence(0) | possibly_damaging(0.477) | 25.2 | 0.32006 | T | T | D |
|  | chr9 | 69205273 | C | A | L38I | | . | | . | . | 2 | tolerated_low_confidence(0.16) | benign(0.034) | 0.507 | 0.00703 | T | T | T |
|  | chr9 | 69216363 | G | T | A24S | | 1 | | 0 | 0 | 2 | deleterious(0) | probably_damaging(0.997) | 29.6 | 0.70427 | T | T | D |
|  | chr9 | 69234509 | G | A | G339D | | 1 | | 1 | 0 | 2 | deleterious(0) | probably_damaging(0.976) | 29.1 | 0.88963 | T | D | D |
|  | chr9 | 69248193 | C | T | T927I | | 1 | | 0 | . | 2 | deleterious(0) | benign(0.022) | 24.8 | 0.22919 | T | T | T |
|  | chr9 | 69254222 | G | C | E1142Q | | 0 | | 0 | . | 2 | deleterious(0.02) | probably_damaging(0.979) | 26.6 | 0.5519 | T | T | D |
|  | chr9 | 69254357 | C | T | R1187W | | 1 | | 1 | . | 2 | deleterious_low_confidence(0) | probably_damaging(1) | 24.8 | 0.49511 | T | T | D |
|  | chr9 | 69216385 | A | G | N31S | | 1 | | 0 | 1 | 2 | deleterious(0.01) | probably_damaging(0.985) | 25.6 | 0.59638 | T | T | D |
|  | chr9 | 69216409 | C | T | T39M | | 1 | | 1 | 1 | 2 | deleterious(0.04) | probably_damaging(0.999) | 25 | 0.40608 | T | T | T |
|  | chr9 | 69216454 | G | C | G54A | | 2 | | 0 | 1 | 2 | deleterious(0) | probably_damaging(0.999) | 27.1 | 0.74021 | T | T | D |
|  | chr9 | 69216456 | C | A | L55M | | 1 | | 0 | 0 | 2 | deleterious(0.04) | probably_damaging(0.999) | 23.2 | 0.36201 | T | T | T |
|  | chr9 | 69218297 | G | A | E71K | | 0 | | 1 | 1 | 2 | deleterious(0) | possibly_damaging(0.658) | 28.7 | 0.59393 | T | T | T |
|  | chr9 | 69218318 | G | T | A78S | | 1 | | 0 | 1 | 2 | deleterious(0.02) | possibly_damaging(0.801) | 27.3 | 0.81298 | T | T | D |
|  | chr9 | 69218351 | G | A | A89T | | 0 | | 0 | 1 | 2 | deleterious(0) | possibly_damaging(0.735) | 28.9 | 0.92601 | T | T | D |
|  | chr9 | 69221403 | C | A | R45S | | 0 | | 0 | . | 2 | deleterious(0.02) | benign(0.165) | 22.1 | 0.32697 | T | T | D |
|  | chr9 | 69225355 | C | T | T93M | | 2 | | 1 | 0 | 2 | deleterious(0.02) | benign(0.286) | 24.5 | 0.5976 | T | T | D |
|  | chr9 | 69227983 | C | T | S199F | | 1 | | 0 | 0 | 2 | deleterious(0.01) | possibly_damaging(0.681) | 24.2 | 0.19911 | T | T | T |
|  | chr9 | 69228083 | C | T,G | N232K | | 0 | | 1 | . | 2 | tolerated(0.81) | benign(0) | 7.87 | 0.06539 | T | T | T |
|  | chr9 | 69236130 | G | T | G386V | | 1 | | 1 | 0 | 2 | deleterious(0) | probably_damaging(1) | 28.4 | 0.88448 | T | T | D |
|  | chr9 | 69237073 | C | T | L464F | | 0 | | 0 | . | 2 | deleterious(0) | probably_damaging(1) | 24.4 | 0.44999 | T | T | D |
|  | chr9 | 69237097 | A | C | T472P | | 1 | | 1 | 0 | 2 | tolerated(0.09) | probably_damaging(0.999) | 26.5 | 0.77141 | T | T | D |
|  | chr9 | 69238758 | G | A | R533Q | | 1 | | 1 | 0 | 2 | deleterious(0) | probably_damaging(0.999) | 32 | 0.80653 | T | T | D |
|  | chr9 | 69239944 | A | T | H546L | | 1 | | 0 | 0 | 2 | deleterious(0) | benign(0.135) | 27.3 | 0.92397 | T | T | D |
|  | chr9 | 69246785 | G | A | G865R | | 1 | | 1 | . | 2 | deleterious(0) | probably_damaging(0.982) | 29.8 | 0.69678 | T | T | T |
|  | chr9 | 69248052 | A | G | Y880C | | 1 | | 1 | . | 2 | deleterious(0.03) | benign(0.106) | 27.2 | 0.62547 | T | T | D |
|  | chr9 | 69251333 | T | G | M1074R | | 1 | | 0 | . | 2 | deleterious_low_confidence(0) | possibly_damaging(0.791) | 25.2 | 0.4655 | T | T | D |

Foot note: Varsome prediction: 0, pathogenic; 1, likely pathogenic; 2, variant of unknown significance; 3, likely benign; 4, benign. Conserved: 0, not conserved; 1, highly conserved. Disease propensity: 0, low disease propensity; 1, high disease propensity. Other variants in region: 0, no; 1, yes. Abbreviations: ALT, alternative; CHR, chromosome; D, deleterious; POS, position; REF, reference; SNV, Single Nucleotide Variant; T, tolerant.

**Supplementary table 2. *Variants identified in volunteers with raised TSBA concentrations but no diagnosis of ICP based on electronic health records.***

| All variants in patients with raised TSBA but no diagnosis of ICP | | | | | |
| --- | --- | --- | --- | --- | --- |
| **Volunteer** | **Gene** | **Variants** | **Zygosity** | **Type** | **Highest BA (umol/L)** |
| 2 | ATP8B1 | M674T | het | Non-synonymous | 13.4 |
| 2 | ATP8B1 | I577V | het | Non-synonymous |  |
| 2 | TJP2 | Q105K | hom | Non-synonymous |  |
| 10 | ABCB4 | T175A | het | Non-synonymous | 11 |
| * 11 | TJP2 | R105K | het | Non-synonymous | 24 |
| 14 | TJP2 | Q105K | het | Non-synonymous | 11.1 |
| # 16 | ABCB11 | M677V | het | Non-synonymous | 14 |
| 17 | ABCB11 | V284A | het | Non-synonymous | 11 |
| * 24 | ABCB11 | N591S | het | Non-synonymous | 343 |
| 24 | ABCB4 | M113V | het | Non-synonymous |  |
| 24 | TJP2 | Q105K | hom | Non-synonymous |  |
| 27 | ABCB11 | N591S | het | Non-synonymous | 10 |
| 28 | NH1R4 | H258R | het | Non-synonymous | 13 |
| 36 | ABCB11 | M677V | hom | Non-synonymous | 11.2 |

Foot note: Abbreviations: Het, heterozygous; hom, homozygous; ICP, intrahepatic cholestasis of pregnancy; TSBA, total serum bile acids. Symbols: #, non-pregnant- history of gallstone, hepatitis C and cirrhosis of liver; *possible diagnosis of ICP. Please note, all other volunteers were identified to be pregnant but to not have a diagnosis of ICP.

**Supplementary table 3. *ABCB4 variants identified with a cholestatic phenotype reported in the literature or no phenotype previously reported.***

| **Clinical phenotype** | **Gene** | **Transcript** | **Protein change** | **dbSNP** | **gnomAD AF** | **G&H AF*** | **ACMG-AMP classification** | **ACMG-AMP criteria** | **Clinvar** | **Ref** |
| --- | --- | --- | --- | --- | --- | --- | --- | --- | --- | --- |
| Cholestatic phenotype reported in the literature | ABCB4 | ENSP00000496956.1:p.Gln1106His | Q1106H ^ | rs779653372 | 0.00001219 | 0.00009889 | LP | PM1, PM2, PP2 | . | [1] |
|  | ABCB4 | ENSP00000496956.1:p.Thr775Met | T775M | rs14805219 | 0.00050410 | 0.00028774 | VUS | PM1, PM2, PP2, BP4 | . | [2-9] |
|  | ABCB4 | ENSP00000496956.1:p.Lys679Arg | K679R | rs374432576 | 0.00000813 | 0.00009568 | VUS | PM2, PP2, BP4 | . | [10] |
|  | ABCB4 | ENSP00000496956.1:p.Arg590Gln | R590Q | rs45575636 | 0.00449100 | 0.00029002 | Benign | PM1, PP2, PP3 | conflicting | [3, 4, 11-17] |
|  | ABCB4 | ENSP00000496956.1:p.Phe522Ser | F522S | . | . | 0.00009575 | LP | PM1, PM2, PP2, PP3 | . | [18] |
|  | ABCB4 | ENSP00000496956.1:p.Tyr467Asp | Y467D | rs1472679087 | 0.00000813 | 0.00009557 | LP | PM1, PM2. PP2 | . | [15] |
|  | ABCB4 | ENSP00000496956.1:p.Gly270Arg | G270R | rs551234479 | 0.00023170 | 0.00057296 | LP | PM1, PM2, PP2, PP3 | VUS | [19] |
|  | ABCB4 | ENSP00000496956.1:p.Met113Ile | M113V | rs752700100 | 0.00005719 | 0.00078705 | LP | PM1, PM2, PP2 | . | [1, 3] |
|  | ABCB4 | ENSP00000496956.1:p.Leu73Val | L73V | rs8187788 | 0.000707 | 0.00009569 | LP | PP2, PP5, BP4 | VUS | [7, 11, 12, 20-22] |
| No phenotype reported in the literature | ABCB4 | ENSP00000496956.1:p.Asn1238Ser | N1238S | . | . | 0.00010739 | VUS | PM1, PM2, PP2, BP4 | . | . |
|  | ABCB4 | ENSP00000496956.1:p.Arg1224His | R1224H | rs144790968 | 0.00003254 | 0.00009804 | LP | PM1, PM2, PP2, PP3 | . | . |
|  | ABCB4 | ENSP00000496956.1:p.Ile1153Lys | I1153K | . | . | 0.00009623 | LP | PM1, PM2, PP2, PP3 | . | . |
|  | ABCB4 | ENSP00000496956.1:p.Cys1124Arg | C1124R | . | . | 0.00009582 | LP | PM1, PM2, PP2, PP3 | . | . |
|  | ABCB4 | ENSP00000496956.1:p.Gly1113Arg | G1113R | rs1282338773 | . | 0.00019562 | LP | PM1, PM2, PP2, PP3 | VUS | . |
|  | ABCB4 | ENSP00000496956.1:p.Val1093Met | V1093M | . | . | 0.00018657 | LP | PM1, PM2, PP2, PP3 | . | . |
|  | ABCB4 | ENSP00000496956.1:p.Lys1060Arg | K1060R | . | . | 0.00028796 | VUS | PM1, PM2, PP2, BP4 | . | . |
|  | ABCB4 | ENSP00000496956.1:p.Ala1004Glu | A1004E | . | . | 0.00009564 | VUS | PM2, PP2, PP3 | . | . |
|  | ABCB4 | ENSP00000496956.1:p.Asp804Asn | D804N | rs762755781 | 0.00000406 | 0.00009590 | LP | PM1, PM2, PP2, PP3 | . | . |
|  | ABCB4 | ENSP00000496956.1:p.Ile753Thr | I753T | . | . | 0.00028719 | VUS | PM1, PM2, PP2, BP4 | . | . |
|  | ABCB4 | ENSP00000496956.1:p.Gln748Glu | Q748E | . | . | 0.00009584 | VUS | PM1, PM2, PP2, BP4 | . | . |
|  | ABCB4 | ENSP00000496956.1:p.Glu684Val | E684V | . | . | 0.00009569 | VUS | PM2, PP2, BP4 | . | . |
|  | ABCB4 | ENSP00000496956.1:p.Trp658Leu | W658L | . | . | 0.00009580 | VUS | PM2, PP2, BP4 | . | . |
|  | ABCB4 | ENSP00000496956.1:p.Ala578Ser | A578S | . | . | 0.00066079 | LP | PM1, PM2, PP2 | . | . |
|  | ABCB4 | ENSP00000496956.1:p.Ile342Thr | I342T | . | . | 0.00009758 | LB | PM1, PM2, PP2 | . | . |
|  | ABCB4 | ENSP00000496956.1:p.Val192Ile | V192I | . | . | 0.00009557 | LP | PM1, PM2, PP2, PP3 | . | . |
|  | ABCB4 | ENSP00000496956.1:p.Arg147Lys | R147K | . | . | 0.00009566 | VUS | PM1, PM2, PP2, BP4 | . | . |
|  | ABCB4 | ENSP00000496956.1:p.Gly124Cys | G124C | . | . | 0.00009569 | LP | PM1, PM2, PM5, PP2, PP3 | . | . |
|  | ABCB4 | ENSP00000496956.1:p.Lys35Glu | K35E | rs879209136 | . | 0.00018748 | VUS | PM2, PP2, BP4 | . | . |

Foot note: Variants were filtered and annotated if they met the following inclusion criteria (MAF < 5%): 1. associated with a phenotype; 2. known in the literature; 3. no recorded GnomAD allele frequency; 4. predicted to be likely pathogenic (LP) based on all 7 *in-silico* predictors. Abbreviations: AF, allele frequency; ACMG-AMP, American College of Medical Genetics and Genomics and the Association for Molecular Pathology (BP, benign supporting; PM, pathogenic moderate; PP, pathogenic supporting); G&H, Genes & Health; LP, likely pathogenic; Ref, references; VUS, variant of unknown significance. Symbols: *Allele frequency specific to East London Genes & Health cohort; ^ see Supplementary Figure 4 for an illustrative example.

**Supplementary table 4. *ABCB11 variants identified with a cholestatic phenotype reported in the literature or no phenotype previously reported.***

| **Clinical Phenotype** | **Gene** | **Transcript** | **Protein change** | **dbSNP** | **gnomAD AF** | **G&H AF*** | **ACMG-AMP classification** | **ACMG-AMP criteria** | **Clinvar** | **Ref** |
| --- | --- | --- | --- | --- | --- | --- | --- | --- | --- | --- |
| Cholestatic phenotype reported in the literature | ABCB11 | ENSP00000497931.1:p.Arg1268Gln | R1268Q | rs72549394 | . | 0.00021983 | LP | PM1, PM2, PP2, PP3, PP5 | . | [23-26] |
|  | ABCB11 | ENSP00000497931.1:p.Arg1153His | R1153H | rs748862206 | 0.00000813 | 0.00009584 | LP | PM1, PM2, PM5, PP2, PP3, PP5 | pathogenic | [25-30] |
|  | ABCB11 | ENSP00000497931.1:p.Arg1050Cys | R1050C | rs72549398 | 0.00001259 | 0.00009562 | LP | PM2, PP2, PP3, PP5 | pathogenic | [23, 27, 31-36] |
|  | ABCB11 | ENSP00000497931.1:p.Ala995Val | A995V | rs868669576 | 0.00000406 | 0.00009551 | LP | PM1, PM2, PP2, PP3 | . | [15] |
|  | ABCB11 | ENSP00000497931.1:p.Arg928Gln | R928Q | rs200488448 | 0.00003257 | 0.00010113 | VUS | PM1, PM2, PP2, BP4 | VUS | [37] |
|  | ABCB11 | ENSP00000497931.1:p.Arg616His | R616H | rs777021400 | 0.00002045 | 0.00009787 | LP | PM1, PM2, PP2, PP3 | VUS | [38] |
|  | ABCB11 | ENSP00000497931.1:p.Thr463Ile | T463I | rs1163486377 | . | 0.00009628 | LP | PM1, PM2, PP2, PP3 | . | [25, 26] |
|  | ABCB11 | ENSP00000497931.1:p.Ser1321Asn | S1321N | rs201693189 | 0.00014620 | 0.00097447 | VUS | PM2, PP2, PP3 | . | [39] |
|  | ABCB11 | ENSP00000497931.1:p.Ala1283Val | A1283V | rs372886308 | 0.00006548 | 0.00009619 | LP | PM1, PM2, PP2, PP3 | VUS | [39] |
|  | ABCB11 | ENSP00000497931.1:p.Asn1211Asp | N1211D | rs762729346 | 0.00006544 | 0.00019110 | VUS | PM1, PM2, PP2 | . | [26] |
|  | ABCB11 | ENSP00000497931.1:p.Ala865Val | A865V | rs118109635 | 0.00108500 | 0.00057361 | Benign | PM1, PP2, PP3, BP6, BS1, BS2 | benign/likely benign | [17, 37, 39-46] |
|  | ABCB11 | ENSP00000497931.1:p.Leu712Ser | L712S | rs372939910 | 0.00002690 | 0.00028852 | VUS | PM1, PP2, BP4 | benign/likely benign | [47] |
|  | ABCB11 | ENSP00000497931.1:p.Glu709Lys | E709K | rs201800225 | 0.00021450 | 0.00019260 | VUS | PM2, PP2, BP4 | VUS | [10, 25] |
|  | ABCB11 | ENSP00000497931.1:p.Arg387His | R387H | rs372784355 | 0.00000813 | 0.00009626 | VUS | PM2, PP2, PP3 | . | [48] |
| No  phenotype reported in the literature | ABCB11 | ENSP00000497931.1:p.Ala1283Ser | A1283S | . | . | 0.00009636 | VUS | PM1, PM2, PP2, BP4 | . | . |
|  | ABCB11 | ENSP00000497931.1:p.Arg1235Gln | R1235Q | rs750033238 | 0.00001218 | 0.00019099 | LP | PM1, PM2, PP2, PP3 | . | . |
|  | ABCB11 | ENSP00000497931.1:p.Asp1172Gly | D1172G | . | . | 0.00009582 | LP | PM1, PM2, PP2, PP3 | . | . |
|  | ABCB11 | ENSP00000497931.1:p.Lys718Glu | K718E | . | . | 0.00009666 | VUS | PM2, PP2, BP4 | . | . |
|  | ABCB11 | ENSP00000497931.1:p.Ser488Cys | S488C | rs777671329 | 0.00000409 | 0.00009549 | LP | PM1, PM2, PP2, PP3 | . | . |
|  | ABCB11 | ENSP00000497931.1:p.Met174Ile | M174I | . | . | 0.00009617 | VUS | PM1, PM2, PP2, BP4 | . | . |
|  | ABCB11 | ENSP00000497931.1:p.Asn116Ser | N116S | . | . | 0.00009569 | LP | PM1, PM2, PP2 | . | . |
|  | ABCB11 | ENSP00000497931.1:p.Pro77Leu | P77L | . | . | 0.00019110 | LP | PM1, PM2, PP2, PP3 | . | . |
|  | ABCB11 | ENSP00000497931.1:p.Ala1287Val | A1287V | . | . | 0.00009601 | VUS | PM1, PM2, PP2 | . | . |
|  | ABCB11 | ENSP00000497931.1:p.Val1212Ile | V1212I | rs546906441 | 0.00003697 | 0.00028664 | LP | PM1, PM2, PP2, PP3 | VUS | . |
|  | ABCB11 | ENSP00000497931.1:p.Thr1180Ala | T1180A | . | . | 0.00009621 | VUS | PM1, PM2, PP2, BP4 | . | . |
|  | ABCB11 | ENSP00000497931.1:p.Gln733Glu | Q733E | . | . | 0.00013839 | VUS | PM2, PP2, BP4 | . | . |
|  | ABCB11 | ENSP00000497931.1:p.Phe651Ser | F651S | rs868080119 | . | 0.00009560 | LP | PM1, PM2, PP2, PP3 | . | . |
|  | ABCB11 | ENSP00000497931.1:p.Arg616Cys | R616C | rs369187042 | 0.00001227 | 0.00009745 | LP | PM1, PM2, PP2, PP3 | VUS | . |
|  | ABCB11 | ENSP00000497931.1:p.Arg313Leu | R313L | . | . | 0.00023585 | VUS | PM1, PM2, PP2, BP4 | . | . |
|  | ABCB11 | ENSP00000497931.1:p.Ile245Phe | I245F | . | . | 0.00009555 | LP | PM1, PM2, PP2, PP3 | . | . |
|  | ABCB11 | ENSP00000497931.1:p.Asp191Ala | D191A | . | . | 0.00009577 | LP | PM1, PM2, PP2, PP3 | . | . |
|  | ABCB11 | ENSP00000497931.1:p.Ile74Val | I74V | . | . | 0.00009555 | VUS | PM1, PM2, PP2, BP4 | . | . |
|  | ABCB11 | ENSP00000497931.1:p.Glu15Asp | E15D | . | . | 0.00009835 | VUS | PM2, PP2, BP4 | . | . |

Foot note: Variants were filtered and annotated if they met the following inclusion criteria (MAF < 5%): 1. associated with a phenotype; 2. known in the literature; 3. no recorded GnomAD allele frequency; 4. predicted to be likely pathogenic (LP) based on all 7 *in-silico* predictors. Abbreviations: AF, allele frequency; ACMG-AMP, American College of Medical Genetics and Genomics and the Association for Molecular Pathology (BP, benign supporting; PM, pathogenic moderate; PP, pathogenic supporting); G&H, Genes & Health; LP, likely pathogenic; Ref, references; VUS, variant of unknown significance. Symbols: *Allele frequency specific to East London Genes & Health cohort.

**Supplementary table 5. *ATP8B1 variants identified with a cholestatic phenotype reported in the literature or no phenotype previously reported.***

| **Clinical phenotype** | **Gene** | **Transcript** | **Protein change** | **dbSNP** | **gnomAD AF*** | **G&H AF*** | **ACMG-AMP classification** | **ACMG-AMP criteria** | **Clinvar** | **Ref** |
| --- | --- | --- | --- | --- | --- | --- | --- | --- | --- | --- |
| Cholestatic phenotype reported in the literature | ATP8B1 | ENSP00000497896.1:p.Ala976Glu | A976E | . | . | 0.00009564 | VUS | PM2, PP2, PP3 | . | [49] |
|  | ATP8B1 | ENSP00000497896.1:p.Arg849Gln | R849Q | rs144656719 | 0.00009342 | 0.00057372 | VUS | PM2, PP2, BP4 | VUS | [50] |
|  | ATP8B1 | ENSP00000497896.1:p.Gly308Ser | G308S | rs1007521320 | 0.00000406 | 0.00009617 | LP | PM2, PM5, PP2, PP3 | . | [51-53] |
|  | ATP8B1 | ENSP00000497896.1:p.Tyr924Cys | Y924C | rs145287364 | 0.00003249 | 0.00009549 | VUS | PM2, PP2, PP3, BP6 | VUS | [49] |
|  | ATP8B1 | ENSP00000497896.1:p.Arg833Gln | R833Q ^1.^ | rs568134011 | 0.00135300 | 0.01042260 | Benign | PM2, PP2, BP4 | Benign | [49] |
|  | ATP8B1 | ENSP00000497896.1:p.Glu429Ala | E429A | rs34018205 | 0.00061740 | 0.00009858 | LB | PM2, PP2, BP4, BP6 | Conflicting | [48, 49, 54, 55] |
|  | ATP8B1 | ENSP00000497896.1:p.Asn45Thr | N45T | rs146599962 | 0.00471500 | 0.00047856 | Benign | PM1, PP2, PP5, BS1, BS2, BP4, BP6 | Likely benign | [3, 7, 39, 49, 56-60] |
| No phenotype reported in the literature | ATP8B1 | ENSP00000497896.1:p.Val868Ile | V868I | rs750735500 | 0.00002031 | 0.00009573 | VUS | PM2, PP2, PP3 | . | . |
|  | ATP8B1 | ENSP00000497896.1:p.Arg768Gly | R768G | . | . | 0.00009810 | VUS | PM2, PP2, BP4 | . | . |
|  | ATP8B1 | ENSP00000497896.1:p.Val730Ala | V730A | rs1302793202 | 0.00000406 | 0.00019146 | VUS | PM2, PP2, PP3 | . | . |
|  | ATP8B1 | ENSP00000497896.1:p.Tyr450Cys | Y450C | . | . | 0.00009741 | VUS | PM2, PP2, PP3 | . | . |
|  | ATP8B1 | ENSP00000497896.1:p.Ala364Glu | A364E | . | . | 0.00009557 | VUS | PM2, PP2, BP4 | . | . |
|  | ATP8B1 | ENSP00000497896.1:p.Gly308Ala | G308A | . | . | 0.00009628 | LP | PM2, PM5, PP2, PP3 | . | . |
|  | ATP8B1 | ENSP00000497896.1:p.Val226Met | V226M | rs1449849281 | 0.00000406 | 0.00010030 | VUS | PM2, PP2, PP3 | . | . |
|  | ATP8B1 | ENSP00000497896.1:p.Ile212Val | I212V | . | . | 0.00028907 | VUS | PM2, PP2, BP4 | . | . |
|  | ATP8B1 | ENSP00000497896.1:p.Met169Thr | M169T | . | . | 0.00009582 | VUS | PM2, PP2 | . | . |
|  | ATP8B1 | ENSP00000497896.1:p.Met110Ile | M110I | . | . | 0.00013214 | VUS | PM1, PM2, PP2, BP4 | . | . |
|  | ATP8B1 | ENSP00000497896.1:p.Cys62Phe | C62F | . | . | 0.00068752 | VUS | PM2, PP2, BP4 | . | . |
|  | ATP8B1 | ENSP00000497896.1:p.Gln1061Leu | Q1061L | . | . | 0.00009549 | VUS | PM2, PP2 | . | . |
|  | ATP8B1 | ENSP00000497896.1:p.Arg797His | R797H | rs776245959 | 0.00001219 | 0.00009566 | VUS | PM2, PP2, BP4 | . | . |
|  | ATP8B1 | ENSP00000497896.1:p.Arg628Gln | R628Q | rs747906077 | 0.00001218 | 0.00009558 | LP | PM2, PM5, PP2, PP3 | . | . |
|  | ATP8B1 | ENSP00000497896.1:p.Leu128Val | L128V | . | . | 0.00012324 | VUS | PM1, PM2, PP2 | . | . |

Foot note: Variants were filtered and annotated if they met the following inclusion criteria (MAF < 5%): 1. associated with a phenotype; 2. known in the literature; 3. no recorded GnomAD allele frequency; 4. predicted to be likely pathogenic (LP) based on all 7 *in-silico* predictors. Abbreviations: AF, allele frequency; ACMG-AMP, American College of Medical Genetics and Genomics and the Association for Molecular Pathology (BP, benign supporting; PM, pathogenic moderate; PP, pathogenic supporting); G&H, Genes & Health; LP, likely pathogenic; Ref, references; VUS, variant of unknown significance. Symbols: *Allele frequency specific to East London Genes & Health cohort; ^1.^ R833Q, Hom (n) 1, Het (n) 110.

**Supplementary table 6. *NR1H4 variants identified with no cholestatic phenotype previously reported.***

| **Clinical phenotype** | **Gene** | **Transcript** | **Protein change** | **dbSNP** | **gnomAD AF*** | **G&H AF*** | **ACMG-AMP classification** | **ACMG-AMP criteria** | **Clinvar** | **Ref** |
| --- | --- | --- | --- | --- | --- | --- | --- | --- | --- | --- |
| No phenotype reported in the literature | NR1H4 | ENSP00000496908.1:p.Asn51Asp | N51D | . | . | 0.00009549 | VUS | PM2, BP4 | . | . |
|  | NR1H4 | ENSP00000496908.1:p.His258Arg | H258R | rs780917996 | . | 0.00038373 | VUS | PM2, BP4 | . | . |
|  | NR1H4 | ENSP00000496908.1:p.Asp316Glu | D316E | rs778153216 | 0.00010970 | 0.00038270 | VUS | PM2, PP3 | . | . |
|  | NR1H4 | ENSP00000496908.1:p.Lys374Glu | K374E | rs906730263 | . | 0.00017100 | VUS | PM2, PP3 | . | . |
|  | NR1H4 | ENSP00000496908.1:p.Pro45Ser | P45S | . | . | 0.00009553 | VUS | PM2, BP4 | . | . |
|  | NR1H4 | ENSP00000496908.1:p.Glu181Asp | E181D | . | . | 0.00009553 | VUS | PM1, PM2, PP3 | . | . |
|  | NR1H4 | ENSP00000496908.1:p.Gln222Leu | Q222L | . | . | 0.00009551 | VUS | PM2 | . | . |

Foot note: Variants were filtered and annotated if they met the following inclusion criteria (MAF < 5%): 1. associated with a phenotype; 2. known in the literature; 3. no recorded GnomAD allele frequency; 4. predicted to be likely pathogenic (LP) based on all 7 *in-silico* predictors. Ref, references. ACMG-AMP, American College of Medical Genetics and Genomics and the Association for Molecular Pathology. Abbreviations: AF, allele frequency; ACMG-AMP, American College of Medical Genetics and Genomics and the Association for Molecular Pathology (BP, benign supporting; PM, pathogenic moderate; PP, pathogenic supporting); G&H, Genes & Health; LP, likely pathogenic; Ref, references; VUS, variant of unknown significance. Symbols: *Allele frequency specific to East London Genes & Health cohort.

**Supplementary table 7. *TJP2 variants identified with a cholestatic phenotype reported in the literature or no phenotype previously reported.***

| **Clinical phenotype** | **Gene** | **Transcript** | **Protein change** | **dbSNP** | **gnomAD AF** | **G&H AF*** | **ACMG-AMP classification** | **ACMG-AMP criteria** | **Clinvar** | **Ref** |
| --- | --- | --- | --- | --- | --- | --- | --- | --- | --- | --- |
| Cholestatic phenotype reported in the literature | TJP2 | ENSP00000497539.1:p.Val3Leu | V3L | rs758381001 | 0.00001343 | 0.00009966 | VUS | PM2, BP4 | . | [7] |
|  | TJP2 | ENSP00000496791.1:p.Thr39Met | T39M | rs138241615 | 0.00147400 | 0.00019309 | VUS | PM2, BP4 | . | [3, 61] |
|  | TJP2 | ENSP00000496791.1:p.Ala89Thr | A89T | rs144396411 | 0.00023970 | 0.00019194 | VUS | PM1, PP3 | Conflicting | [62] |
|  | TJP2 | ENSP00000497787.1:p.Thr93Met | T93M | rs548602675 | 0.00000813 | 0.00009579 | VUS | PM2, PP3 | . | [7] |
| No phenotype reported in the literature | TJP2 | ENSP00000496791.1:p.Gly865Arg | G865R | . | . | 0.00009558 | VUS | PM2, PP3 | . | . |
|  | TJP2 | ENSP00000438262.1:p.Leu38Ile | L38I | . | . | 0.00009643 | VUS | PM2, BP4 | . | . |
|  | TJP2 | ENSP00000496791.1:p.Ala24Ser | A24S | . | . | 0.00009568 | VUS | PM1, PM2, PP3 | . | . |
|  | TJP2 | ENSP00000497787.1:p.Gly339Asp | G339D | . | . | 0.00011364 | VUS | PM1, PM2, PP3 | . | . |
|  | TJP2 | ENSP00000496791.1:p.Thr927Ile | T927I | . | . | 0.00009677 | VUS | PM2, BP4 | . | . |
|  | TJP2 | ENSP00000497861.1:p.Glu1142Gln | E1142Q | . | . | 0.00009558 | VUS | PM1, PM2, PP3 | . | . |
|  | TJP2 | ENSP00000497861.1:p.Arg1187Trp | R1187W | rs112109886 | . | 0.00009968 | VUS | PM1, PM2, PP3 | . | . |
|  | TJP2 | ENSP00000496791.1:p.Asn31Ser | N31S | . | . | 0.00009579 | VUS | PM1, PM2, PP3 | . | . |
|  | TJP2 | ENSP00000496791.1:p.Gly54Ala | G54A | . | . | 0.00011756 | VUS | PM1, PM2, PP3 | . | . |
|  | TJP2 | ENSP00000496791.1:p.Leu55Met | L55M | . | . | 0.00023502 | VUS | PM1, PM2, BP4 | . | . |
|  | TJP2 | ENSP00000496791.1:p.Glu71Lys | E71K | . | . | 0.00009582 | VUS | PM1, PM2 | . | . |
|  | TJP2 | ENSP00000496791.1:p.Ala78Ser | A78S | . | . | 0.00019205 | VUS | PM1, PM2, PP3 | . | . |
|  | TJP2 | ENSP00000497787.1:p.Arg45Ser | R45S | . | . | 0.00009584 | VUS | PM2, BP4 | . | . |
|  | TJP2 | ENSP00000497787.1:p.Ser199Phe | S199F | . | . | 0.00009766 | VUS | PM2 | . | . |
|  | TJP2 | ENSP00000497787.1:p.Asn232Lys | N232K | . | . | 0.00019164 | VUS | PM2, BP4 | . | . |
|  | TJP2 | ENSP00000497787.1:p.Gly386Val | G386V | . | . | 0.00009562 | VUS | PM2, PP3 | . | . |
|  | TJP2 | ENSP00000497787.1:p.Leu464Phe | L464F | . | . | 0.00009817 | VUS | PM2, PP3 | . | . |
|  | TJP2 | ENSP00000497787.1:p.Thr472Pro | T472P | . | . | 0.00010119 | VUS | PM2, PP3 | . | . |
|  | TJP2 | ENSP00000497787.1:p.Arg533Gln | R533Q | . | . | 0.00009562 | VUS | PM2, PP3 | . | . |
|  | TJP2 | ENSP00000497787.1:p.His546Tyr | H546L | rs760920622 | . | 0.00038197 | VUS | PM2, PP3 | . | . |
|  | TJP2 | ENSP00000496791.1:p.Tyr880Cys | Y880C | . | . | 0.00019593 | VUS | PM2, PP3 | . | . |
|  | TJP2 | ENSP00000496791.1:p.Met1074Arg | M1074R | . | . | 0.00009566 | VUS | PM2 | . | . |

Foot note: Variants were filtered and annotated if they met the following inclusion criteria (MAF < 5%): 1. associated with a phenotype; 2. known in the literature; 3. no recorded GnomAD allele frequency; 4. predicted to be likely pathogenic (LP) based on all 7 *in-silico* predictors. Abbreviations: AF, allele frequency; ACMG-AMP, American College of Medical Genetics and Genomics and the Association for Molecular Pathology (BP, benign supporting; PM, pathogenic moderate; PP, pathogenic supporting); G&H, Genes & Health; LP, likely pathogenic; Ref, references; VUS, variant of unknown significance. Symbols: *Allele frequency specific to East London Genes & Health cohort.

**Supplementary table 8. *Summary of protein modelling results in variants that passed inclusion criteria and had a resolved 3D structure.***

| **Gene** | **Clinical phenotype** | **Variant** | **Features of substitution site** | **CUPSAT** | **∆∆G DUET** | **∆∆G SDM** | **∆∆G mCSM** | **Dynamut** | **Depth (Å)** | **SNPMuSiC** |
| --- | --- | --- | --- | --- | --- | --- | --- | --- | --- | --- |
| ABCB4 | ICP | G1254S | Unresolved 3D structure region |  |  |  |  |  |  |  |
| ABCB4 | ICP | P1050S | Nucleotide binding domain 2 | destabilising, unfavourable torsion angle of Ser residue, ∆∆G -1.53kcal/mol | destabilising -0.066kcal/mol | stabilising, 0.760kcal/mol | destabilising, -0.691 kcal/mol | decrease in molecule flexibility ΔΔSVib ENCoM:  -0.164kcal/mol-1/K-1 | WT (3.5Å) to Mutant (4.0Å) | deleterious effect (0.6) thermodynamic stability - destabilising thermal stability - destabilising |
| ABCB4 | ICP | A833T | Transmembrane domain 2 | destabilising, unfavourable torsion angle of Thr residue, ∆∆G -0.83kcal/mol | destabilising -1.661kcal/mol | destabilising, -2.620kcal/mol | destabilising, -1.487 kcal/mol | decrease in molecule flexibility ΔΔSVib ENCoM:  -0.191kcal/mol-1/K-1 | WT (5.5Å) to Mutant (6.2Å) | neutral effect (-0.45) |
| ABCB4 | ICP | N510S | Nucleotide binding domain 1 | destabilising, unfavourable torsion angle of Ser residue, ∆∆G -0.63kcal/mol | destabilising -0.816kcal/mol | destabilising, -1.300kcal/mol | destabilising -0.688kcal/mol | increase in molecule flexibility  ΔΔSVib ENCoM: 0.361kcal/mol-1/K-1 | WT (3.6Å) to Mutant (3.3Å) | deleterious effect (0.1) thermodynamic stability - destabilising thermal stability - destabilising |
| ABCB4 | Gallstone disease | R1137Q | Nucleotide binding domain 2 | destabilising, favourable torsion angle of Gln residue, ∆∆G -0.21kcal/mol | destabilising -0.261kcal/mol | destabilising, -0.560kcal/mol | destabilising -0.194kcal/mol | increase in molecule flexibility  ΔΔSVib ENCoM: 0.076kcal/mol-1/K-1 | WT (3.5Å) to Mutant (3.5Å) | neutral effect (-0.55) |
| ABCB4 | Gallstone disease | G826R | Transmembrane domain 2 | destabilising, favourable torsion angle of Arg residue, ∆∆G -1.64kcal/mol | destabilising -0.904kcal/mol | destabilising, -2.250kcal/mol | destabilising -0.897kcal/mol | Decrease in molecule flexibility  ΔΔSVib ENCoM: -1.520kcal/mol-1/K-1 | WT (6.4Å) to Mutant (5.5Å) | deleterious effect (0.58) thermodynamic stability - destabilising thermal stability - destabilising |
| ABCB4 | Gallstone disease | R788L | Transmembrane domain 2 | stabilising, favourable torsion angle of Leu residue,  ∆∆G 0.21kcal/mol | destabilising -0.552kcal/mol | stabilising, 0.930kcal/mol | destabilising -1.115kcal/mol | increase in molecule flexibility  ΔΔSVib ENCoM: 0.137kcal/mol-1/K-1 | WT (5.8Å) to Mutant (5.9Å) | deleterious effect (0.57) thermodynamic stability - stabilising thermal stability - stabilising |
| ABCB4 | Gallstone disease | D686N | Unresolved 3D structure region |  |  |  |  |  |  |  |
| ABCB4 | Gallstone disease | M676I | Unresolved 3D structure region |  |  |  |  |  |  |  |
| ABCB4 | Gallstone disease | T651N | Unresolved 3D structure region |  |  |  |  |  |  |  |
| ABCB4 | Gallstone disease | K391E | Nucleotide binding domain 1 | stabilising, unfavourable torsion angle of Glu residue,  ∆∆G 0.61kcal/mol | stabilising, 0.634kcal/mol | stabilising, 0.500kcal/mol | stabilising, 0.244kcal/mol | increase in molecule flexibility  ΔΔSVib ENCoM: 0.135kcal/mol-1/K-1 | WT (3.4Å) to Mutant (3.6Å) | neutral effect (-0.40) |
| ABCB4 | Cholangiocarcinoma | Q668H | Unresolved 3D structure region |  |  |  |  |  |  |  |
| ABCB4 | . | Q1106H | Nucleotide binding domain 2 | destabilising, no change of torsion angle of His residue,  ∆∆G -2.16kcal/mol | destabilising -0.672kcal/mol | stabilising, -0.060kcal/mol | destabilising -0.863kcal/mol | increase in molecule flexibility  ΔΔSVib ENCoM: 0.412kcal/mol-1/K-1 | WT (3.8Å) to Mutant (4.0Å) | neutral effect (-0.24) |
| ABCB4 | . | F522S | Nucleotide binding domain 1 | destabilising, unfavourable torsion angle of Ser residue, ∆∆G -1.06kcal/mol | destabilising -1.486kcal/mol | destabilising, -1.190kcal/mol | destabilising -1.372kcal/mol | increase in molecule flexibility ΔΔSVib ENCoM:  1.320kcal/mol-1/K-1 | WT (4.2Å) to Mutant (3.4Å) | deleterious effect (0.54) thermodynamic stability - destabilising thermal stability - destabilising |
| ABCB4 | . | Y467D | Nucleotide binding domain 1 | destabilising, favourable torsion angle of Asp residue, ∆∆G -1.95kcal/mol | destabilising -1.536kcal/mol | destabilising -0.660kcal/mol | destabilising -1.731kcal/mol | increase in molecule flexibility ΔΔSVib ENCoM:  0.852kcal/mol-1/K-1 | WT (4.2Å) to Mutant (4.4Å) | deleterious effect (0.59) thermodynamic stability - destabilising thermal stability - destabilising |
| ABCB4 | . | G270R | Transmembrane domain 1 | destabilising, favourable torsion angle of Arg residue, ∆∆G -1.91kcal/mol | destabilising -1.296kcal/mol | destabilising -3.150kcal/mol | destabilising -1.045kcal/mol | decrease in molecule flexibility ΔΔSVib ENCoM:  -1.221kcal/mol-1/K-1 | WT (5.3Å) to Mutant (4.3Å) | deleterious effect (0.54) thermodynamic stability - destabilising thermal stability - destabilising |
| ABCB4 | . | G124C | Transmembrane domain 1 | stabilising,  unfavourable torsion angle of Cys residue, ∆∆G 0.66kcal/mol | destabilising -0.745kcal/mol | destabilising -0.560kcal/mol | destabilising -0.745kcal/mol | decrease in molecule flexibility ΔΔSVib ENCoM:  -0.736kcal/mol-1/K-1 | WT (6.0Å) to Mutant (6.3Å) | deleterious effect (0.60) thermodynamic stability - destabilising thermal stability - destabilising |
| ABCB4 | . | M113V | Transmembrane domain 1 | destabilising,  unfavourable torsion angle of Val residue, ∆∆G -2.9kcal/mol | destabilising -1.212kcal/mol | destabilising -1.060kcal/mol | destabilising -1.412kcal/mol | increase in molecule flexibility ΔΔSVib ENCoM:  0.432kcal/mol-1/K-1 | WT (7.5Å) to Mutant (8.1Å) | neutral effect (-0.11) |
| ABCB4 | . | L73V | Transmembrane domain 1 | destabilising,  unfavourable torsion angle of Val residue, ∆∆G -1.61kcal/mol | destabilising -0.478kcal/mol | destabilising -0.590kcal/mol | destabilising -0.709kcal/mol | increase in molecule flexibility ΔΔSVib ENCoM:  0.066kcal/mol-1/K-1 | WT (3.5Å) to Mutant (3.6Å) | neutral effect (-0.46) |
| ABCB4 | . | I1153K | Nucleotide binding domain 2 | stabilising,  favourable torsion angle of Lys residue, ∆∆G 2.69kcal/mol | destabilising -2.074kcal/mol | destabilising -2.210kcal/mol | destabilising -1.884kcal/mol | decrease in molecule flexibility ΔΔSVib ENCoM:  -0.039kcal/mol-1/K-1 | WT (7.4Å) to Mutant (7.6Å) | deleterious effect (0.62) thermodynamic stability - destabilising thermal stability - destabilising |
| ABCB4 | . | C1124R | Nucleotide binding domain 2 | stabilising,  unfavourable torsion angle of Arg residue, ∆∆G 3.44kcal/mol | stabilising, 0.051kcal/mol | stabilising, 0.010kcal/mol | destabilising -0.276kcal/mol | decrease in molecule flexibility ΔΔSVib ENCoM:  -0.074kcal/mol-1/K-1 | WT (3.7Å) to Mutant (4.1Å) | deleterious effect (0.82) thermodynamic stability - destabilising thermal stability - destabilising |
| ABCB4 | . | G1113R | Nucleotide binding domain 2 | stabilising,  favourable torsion angle of Arg residue, ∆∆G 0.52kcal/mol | destabilising -1.123kcal/mol | destabilising -2.140kcal/mol | destabilising -1.114kcal/mol | decrease in molecule flexibility ΔΔSVib ENCoM:  -1.012kcal/mol-1/K-1 | WT (9.8Å) to Mutant (8.9Å) | deleterious effect (0.66) thermodynamic stability - destabilising thermal stability - destabilising |
| ABCB4 | . | V1093M | Nucleotide binding domain 2 | destabilising,  favourable torsion angle of Met residue, ∆∆G -3.29kcal/mol | destabilising -0.963kcal/mol | destabilising -2.390kcal/mol | destabilising -0.586kcal/mol | decrease in molecule flexibility ΔΔSVib ENCoM:  -0.435kcal/mol-1/K-1 | WT (6.3Å) to Mutant (6.6Å) | deleterious effect (0.02) thermodynamic stability - destabilising thermal stability - destabilising |
| ABCB4 | . | A578S | Nucleotide binding domain 1 | destabilising,  unfavourable torsion angle of Ser residue, ∆∆G -2.82kcal/mol | destabilising -0.902kcal/mol | destabilising -1.980kcal/mol | destabilising -0.862kcal/mol | decrease in molecule flexibility ΔΔSVib ENCoM:  -0.277kcal/mol-1/K-1 | WT (6.2Å) to Mutant (6.1Å) | neutral effect (-0.22) |
| ABCB4 | . | V192I | Transmembrane domain 1 | stabilising,  unfavourable torsion angle of Ile residue, ∆∆G 1.12kcal/mol | stabilising, 0.713kcal/mol | destabilising -0.190kcal/mol | stabilising, 0.331kcal/mol | decrease in molecule flexibility ΔΔSVib ENCoM:  -0.123kcal/mol-1/K-1 | WT (5Å) to Mutant (5Å) | neutral effect (-0.44) |
| ABCB4 | . | R1224H | Nucleotide binding domain 2 | stabilising,  favourable torsion angle of His residue, ∆∆G 0.04kcal/mol | destabilising -1.706kcal/mol | destabilising -0.520kcal/mol | destabilising -1.473kcal/mol | decrease in molecule flexibility ΔΔSVib ENCoM:  -0.217kcal/mol-1/K-1 | WT (5.3Å) to Mutant (5.1Å) | deleterious effect (0.13) thermodynamic stability - destabilising thermal stability - destabilising |
| ABCB4 | . | D804N | Transmembrane domain 2 | destabilising,  favourable torsion angle of Asn residue,  ∆∆G -0.79kcal/mol | destabilising -0.078kcal/mol | stabilising, 0.410kcal/mol | destabilising -0.389kcal/mol | increase in molecule flexibility ΔΔSVib ENCoM:  0.074kcal/mol-1/K-1 |  | neutral effect (-0.12) |
| ABCB11 | ICP | ABCB11 D1284N | Nucleotide binding domain 2 | destabilising, favourable torsion angle of Asn residue,  ∆∆G -1.32kcal/mol | stabilising 0.035kcal/mol | stabilising 0.150kcal/mol | destabilising -0.231kcal/mol | decrease in molecule flexibility  ΔΔSVib ENCoM:  -0.190kcal/mol-1/K-1 | WT (4.2Å) to Mutant (4.3Å) | deleterious effect (0.13) thermodynamic stability - destabilising thermal stability - stabilising |
| ABCB11 | ICP | ABCB11 R1050H | Nucleotide binding domain 2 | destabilising, unfavourable torsion angle of His residue,  ∆∆G -0.57kcal/mol | destabilising -1.256kcal/mol | stabilising 0.045kcal/mol | destabilising -1.568kcal/mol | increase in molecule flexibility  ΔΔSVib ENCoM:  0.148kcal/mol-1/K-1 | WT (4.2Å) to Mutant (4.5Å) | neutral effect (-0.62) |
| ABCB11 | ICP | M677V | Unresolved 3D structure region |  |  |  |  |  |  |  |
| ABCB11 | ICP | N591S | Nucleotide binding domain 1 | destabilising, favourable torsion angle of Ser residue,  ∆∆G -1.52kcal/mol | stabilising 0.157kcal/mol | destabilising -0.800kcal/mol | destabilising -0.009kcal/mol | increase in molecule flexibility  ΔΔSVib ENCoM:  0.002kcal/mol-1/K-1 | WT (3.2Å) to Mutant (3.1Å) | neutral effect (-0.50) |
| ABCB11 | ICP | ABCB11 V284A | Transmembrane domain 1 | destabilising, unfavourable torsion angle of Ala residue,  ∆∆G -2.32kcal/mol | stabilising 2.154kcal/mol | destabilising -1.080kcal/mol | destabilising -1.937kcal/mol | increase in molecule flexibility  ΔΔSVib ENCoM:  0.660kcal/mol-1/K-1 | WT (8.4Å) to Mutant (8.5Å) | deleterious effect (0.14) thermodynamic stability - destabilising thermal stability - destabilising |
| ABCB11 | Gallstone disease | A1260P | Nucleotide binding domain 2 | destabilising, unfavourable torsion angle of Pro residue,  ∆∆G -3.32kcal/mol | destabilising -1.328kcal/mol | destabilising -4.450kcal/mol | destabilising -0.651kcal/mol | decrease in molecule flexibility  ΔΔSVib ENCoM:  -0.279kcal/mol-1/K-1 | WT (7.4Å) to Mutant (6.3Å) | deleterious effect (0.46) thermodynamic stability - destabilising thermal stability - destabilising |
| ABCB11 | Gallstone disease | Q976R | Transmembrane domain 2 | destabilising, favourable torsion angle of Arg residue,  ∆∆G -0.33kcal/mol | stabilising 0.240kcal/mol | stabilising 0.380kcal/mol | destabilising -0.098kcal/mol | increase in molecule flexibility  ΔΔSVib ENCoM:  0.150kcal/mol-1/K-1 | WT (3.6Å) to Mutant (3.6Å) | neutral effect (-0.58) |
| ABCB11 | Gallstone disease | A926S | Transmembrane domain 2 | stabilising, favourable torsion angle of Ser residue,  ∆∆G 0.73kcal/mol | destabilising -0.573kcal/mol | destabilising -2.040kcal/mol | destabilising -0.568kcal/mol | decrease in molecule flexibility  ΔΔSVib ENCoM:  -0.076kcal/mol-1/K-1 | WT (3.1Å) to Mutant (3.2Å) | neutral effect (-0.58) |
| ABCB11 | Gallstone disease | A679V | Unresolved 3D structure region |  |  |  |  |  |  |  |
| ABCB11 | Gallstone disease | N539D | Nucleotide binding domain 1 | destabilising, favourable torsion angle of Asp residue,  ∆∆G -0.13kcal/mol | stabilising 0.215kcal/mol | stabilising 0.350kcal/mol | destabilising -0.142kcal/mol | increase in molecule flexibility  ΔΔSVib ENCoM:  0.066kcal/mol-1/K-1 | WT (3.4Å) to Mutant (3.4Å) | neutral effect (-0.51) |
| ABCB11 | Gallstone disease | R487C | Nucleotide binding domain 1 | stabilising, favourable torsion angle of Cys residue,  ∆∆G 2.17kcal/mol | destabilising -0.413kcal/mol | destabilising -0.130kcal/mol | destabilising -0.432kcal/mol | increase in molecule flexibility  ΔΔSVib ENCoM:  0.297kcal/mol-1/K-1 | WT (3.6Å) to Mutant (3.6Å) | deleterious effect (0.44) thermodynamic stability - destabilising thermal stability - destabilising |
| ABCB11 | Gallstone disease | A311T | Transmembrane domain 1 | destabilising, favourable torsion angle of Thr residue,  ∆∆G -0.87kcal/mol | destabilising -1.905kcal/mol | destabilising -2.540kcal/mol | destabilising -1.769kcal/mol | decrease in molecule flexibility  ΔΔSVib ENCoM:  -0.156kcal/mol-1/K-1 | WT (7.7Å) to Mutant (8.6Å) | neutral effect (-0.07) |
| ABCB11 | Gallstone disease | V95I | Transmembrane domain 1 | destabilising, unfavourable torsion angle of Ile residue,  ∆∆G -0.27kcal/mol | destabilising -0.172kcal/mol | stabilising 0.360kcal/mol | destabilising -0.647kcal/mol | decrease in molecule flexibility  ΔΔSVib ENCoM:  -0.250kcal/mol-1/K-1 | WT (3.6Å) to Mutant (3.8Å) | neutral effect (-0.67) |
| ABCB11 | Gallstone disease | D94N | Transmembrane domain 1 | destabilising, unfavourable torsion angle of Asn residue,  ∆∆G -1.59kcal/mol | destabilising -0.477kcal/mol | stabilising 0.570kcal/mol | destabilising -0.884kcal/mol | increase in molecule flexibility  ΔΔSVib ENCoM:  0.180kcal/mol-1/K-1 | WT (5.3Å) to Mutant (6.0Å) | neutral effect (-0.03) |
| ABCB11 | Gallstone disease | K12R | Unresolved 3D structure region |  |  |  |  |  |  |  |
| ABCB11 | . | R1268Q | Nucleotide binding domain 2 | destabilising, unfavourable torsion angle of Gln residue,  ∆∆G -1.21kcal/mol | destabilising -0.578kcal/mol | destabilising -0.750kcal/mol | destabilising -0.614kcal/mol | increase in molecule flexibility  ΔΔSVib ENCoM:  0.478kcal/mol-1/K-1 | WT (4.4Å) to Mutant (4.5Å) | deleterious effect (0.01) thermodynamic stability - destabilising thermal stability - destabilising |
| ABCB11 | . | R1153H | Nucleotide binding domain 2 | destabilising, unfavourable torsion angle of His residue,  ∆∆G -1.21kcal/mol | destabilising -2.355kcal/mol | destabilising -0.560kcal/mol | destabilising -2.156kcal/mol | increase in molecule flexibility  ΔΔSVib ENCoM:  0.014kcal/mol-1/K-1 | WT (5.3Å) to Mutant (5.4Å) | deleterious effect (0.10) thermodynamic stability - destabilising thermal stability - destabilising |
| ABCB11 | . | R1050C | Nucleotide binding domain 2 | destabilising, unfavourable torsion angle of Cys residue,  ∆∆G -0.06kcal/mol | destabilising -0.986kcal/mol | destabilising -0.660kcal/mol | destabilising -0.988kcal/mol | increase in molecule flexibility  ΔΔSVib ENCoM:  0.227kcal/mol-1/K-1 | WT (4.2Å) to Mutant (4.2Å) | deleterious effect (0.16) thermodynamic stability - destabilising thermal stability - destabilising |
| ABCB11 | . | A995V | Transmembrane domain 2 | stabilising, unfavourable torsion angle of Val residue,  ∆∆G 1.74kcal/mol | stabilising 0.474kcal/mol | destabilising -1.030kcal/mol | stabilising 0.291kcal/mol | decrease in molecule flexibility  ΔΔSVib ENCoM:  -0.389kcal/mol-1/K-1 | WT (7.9Å) to Mutant (8.5Å) | deleterious effect (0.19) thermodynamic stability - destabilising thermal stability - destabilising |
| ABCB11 | . | R616H | Nucleotide binding domain 1 | destabilising, unfavourable torsion angle of His residue,  ∆∆G -1.93kcal/mol | destabilising -1.069kcal/mol | stabilising 0.130kcal/mol | destabilising -1.056kcal/mol | decrease in molecule flexibility  ΔΔSVib ENCoM:  -0.977kcal/mol-1/K-1 | WT (3.6Å) to Mutant (3.5Å) | neutral effect (-0.39) |
| ABCB11 | . | T463I | Nucleotide binding domain 1 | stabilising, favourable torsion angle of Ile residue,  ∆∆G 4.64kcal/mol | stabilising 0.642kcal/mol | stabilising 1.030kcal/mol | stabilising 0.054kcal/mol | decrease in molecule flexibility  ΔΔSVib ENCoM:  -0.002kcal/mol-1/K-1 | WT (6.9Å) to Mutant (6.6Å) | deleterious effect (0.51) thermodynamic stability - destabilising thermal stability - destabilising |
| ABCB11 | . | R1235Q | Nucleotide binding domain 2 | destabilising, favourable torsion angle of Gln residue,  ∆∆G -0.69kcal/mol | destabilising -0.920kcal/mol | destabilising -0.750kcal/mol | destabilising -0.809kcal/mol | increase in molecule flexibility  ΔΔSVib ENCoM:  1.025kcal/mol-1/K-1 | WT (5.5Å) to Mutant (4.6Å) | neutral effect (-0.07) |
| ABCB11 | . | D1172G | Nucleotide binding domain 2 | destabilising, unfavourable torsion angle of Arg residue,  ∆∆G -3.01kcal/mol | destabilising -0.260kcal/mol | stabilising 0.020kcal/mol | destabilising -0.553kcal/mol | increase in molecule flexibility  ΔΔSVib ENCoM:  0.277kcal/mol-1/K-1 | WT (3.8Å) to Mutant (4.3Å) | neutral effect (-0.22) |
| ABCB11 | . | S488C | Nucleotide binding domain 1 | stabilising, unfavourable torsion angle of Cys residue,  ∆∆G 1.72kcal/mol | destabilising -0.002kcal/mol | stabilising 1.200kcal/mol | destabilising -0.392kcal/mol | increase in molecule flexibility  ΔΔSVib ENCoM:  0.012kcal/mol-1/K-1 | WT (3.1Å) to Mutant (3.2Å) | neutral effect (-0.15) |
| ABCB11 | . | N116S | Transmembrane domain 1 | stabilising, unfavourable torsion angle of Ser residue,  ∆∆G 1.12kcal/mol | destabilising -0.471kcal/mol | destabilising -1.620kcal/mol | destabilising -0.252kcal/mol | increase in molecule flexibility  ΔΔSVib ENCoM:  0.000kcal/mol-1/K-1 | WT (4.3Å) to Mutant (4.4Å) | neutral effect (-0.03) |
| ABCB11 | . | P77L | Transmembrane domain 1 | destabilising, unfavourable torsion angle of Leu residue,  ∆∆G -4.79kcal/mol | stabilising 0.042kcal/mol | stabilising 2.790kcal/mol | destabilising -0.860kcal/mol | decrease in molecule flexibility  ΔΔSVib ENCoM:  -0.928kcal/mol-1/K-1 | WT (7.3Å) to Mutant (7.3Å) | deleterious effect (0.93) thermodynamic stability - stabilising thermal stability - destabilising |
| ABCB11 | . | V1212I | Nucleotide binding domain 2 | destabilising, unfavourable torsion angle of Ile residue,  ∆∆G -2.73kcal/mol | destabilising -0.367kcal/mol | destabilising -0.180kcal/mol | destabilising -0.631kcal/mol | decrease in molecule flexibility  ΔΔSVib ENCoM:  -0.465kcal/mol-1/K-1 | WT (6.2Å) to Mutant (5.9Å) | neutral effect (-0.15) |
| ABCB11 | . | F651S | Nucleotide binding domain 1 | destabilising, favourable torsion angle of Ser residue,  ∆∆G -0.27kcal/mol | destabilising -2.245kcal/mol | destabilising -1.940kcal/mol | destabilising -2.011kcal/mol | increase in molecule flexibility  ΔΔSVib ENCoM:  0.758kcal/mol-1/K-1 | WT (4.1Å) to Mutant (3.9Å) | deleterious effect (0.33) thermodynamic stability - destabilising thermal stability - destabilising |
| ABCB11 | . | I245F | Transmembrane domain 1 | stabilising, unfavourable torsion angle of Phe residue,  ∆∆G 0.39kcal/mol | destabilising -1.160kcal/mol | destabilising -0.210kcal/mol | destabilising -1.121kcal/mol | decrease in molecule flexibility  ΔΔSVib ENCoM:  -0.461kcal/mol-1/K-1 | WT (7.0Å) to Mutant (6.9Å) | deleterious effect (0.02) thermodynamic stability - destabilising thermal stability - destabilising |
| ABCB11 | . | D191A | Transmembrane domain 1 | destabilising, favourable torsion angle of Ala residue,  ∆∆G -0.55kcal/mol | stabilising 0.089kcal/mol | stabilising 1.300kcal/mol | destabilising -0.495kcal/mol | increase in molecule flexibility  ΔΔSVib ENCoM:  0.395kcal/mol-1/K-1 | WT (4.0Å) to Mutant (3.9Å) | deleterious effect (0.16) thermodynamic stability - destabilising thermal stability - destabilising |
| ATP8B1 | ICP | R384H | Transmembrane domain 1 | stabilising, unfavourable torsion angle of His residue,  ∆∆G 0.54kcal/mol | destabilising -1.573kcal/mol | stabilising 0.49kcal/mol | destabilising -1.915kcal/mol | decrease in molecule flexibility  ΔΔSVib ENCoM:  -0.91kcal/mol-1/K-1 | WT (4.1Å) to Mutant (4.5Å) | neutral effect (-0.29) |
| ATP8B1 | Gallstone disease | V1161A | Transmembrane domain 2 | destabilising, favourable torsion angle of Ala residue,  ∆∆G -3.54kcal/mol | destabilising -1.436kcal/mol | destabilising -0.25kcal/mol | destabilising -1.667kcal/mol | decrease in molecule flexibility  ΔΔSVib ENCoM:  -0.82kcal/mol-1/K-1 | WT (3.4Å) to Mutant (3.2Å) | deleterious effect (0.18) thermodynamic stability - destabilising thermal stability - destabilising |
| ATP8B1 | Gallstone disease | T1092I | Transmembrane domain 2 | destabilising, unfavourable torsion angle of Ile residue,  ∆∆G -2.54kcal/mol | stabilising 0.358kcal/mol | stabilising 1.04kcal/mol | destabilising -0.06kcal/mol | increase in molecule flexibility  ΔΔSVib ENCoM:  0.24kcal/mol-1/K-1 | WT (6.9Å) to Mutant (6.7Å) | deleterious effect (0.41) thermodynamic stability - destabilising thermal stability - destabilising |
| ATP8B1 | Gallstone disease | M674T | Nucleotide binding domain | destabilising, unfavourable torsion angle of Thr residue,  ∆∆G -0.18kcal/mol | stabilising 0.267kcal/mol | destabilising -1.0kcal/mol | destabilising -0.097kcal/mol | increase in molecule flexibility  ΔΔSVib ENCoM:  0.1kcal/mol-1/K-1 | WT (3.3Å) to Mutant (3.2Å) | neutral effect (-0.55) |
| ATP8B1 | Gallstone disease | I577V | Nucleotide binding domain | destabilising, favourable torsion angle of Val residue,  ∆∆G -0.8kcal/mol | destabilising -1.606kcal/mol | destabilising -2.38kcal/mol | destabilising -1.321kcal/mol | decrease in molecule flexibility  ΔΔSVib ENCoM:  -0.88kcal/mol-1/K-1 | WT (9.2Å) to Mutant (9.1Å) | neutral effect (-0.34) |
| ATP8B1 | Gallstone disease | H78Q | Transmembrane domain 1 |  |  |  |  |  |  |  |
| ATP8B1 | Gallstone disease | D14Y | Transmembrane domain 1 |  |  |  |  |  |  |  |
| ATP8B1 | Cirrhosis | I513T | Nucleotide binding domain | destabilising, unfavourable torsion angle of Thr residue,  ∆∆G -2.26kcal/mol | destabilising -3.232kcal/mol | destabilising -2.18kcal/mol | destabilising -3.088cal/mol | decrease in molecule flexibility  ΔΔSVib ENCoM:  -2.98kcal/mol-1/K-1 | WT (5.5Å) to Mutant (4.8Å) | deleterious effect (0.01) thermodynamic stability - destabilising thermal stability - destabilising |
| ATP8B1 | Cirrhosis, secondary malignant neoplasm of liver and bile duct, gallstone disease | D70N | Transmembrane domain 1 | destabilising, favourable torsion angle of Asn residue,  ∆∆G -0.61kcal/mol | stabilising 0.505kcal/mol | stabilising 0.01kcal/mol | stabilising 0.385kcal/mol | increase in molecule flexibility  ΔΔSVib ENCoM:  0.37kcal/mol-1/K-1 | WT (3.4Å) to Mutant (3.5Å) | neutral effect (-0.44) |
| ATP8B1 | . | G308S | Actuator domain | destabilising, favourable torsion angle of Ser residue,  ∆∆G -2.09kcal/mol | destabilising -1.593kcal/mol | destabilising -2.5kcal/mol | destabilising -1.712kcal/mol | decrease in molecule flexibility  ΔΔSVib ENCoM:  -1.4kcal/mol-1/K-1 | WT (12.6Å) to Mutant (12.1Å) | deleterious effect (0.30) thermodynamic stability - destabilising thermal stability - destabilising |
| ATP8B1 | . | G308A | Actuator domain 1 | destabilising, unfavourable torsion angle of Ala residue,  ∆∆G -1.57kcal/mol | destabilising -0.215kcal/mol | destabilising -0.47kcal/mol | destabilising -0.51kcal/mol | decrease in molecule flexibility  ΔΔSVib ENCoM:  -0.09kcal/mol-1/K-1 | WT (12.6Å) to Mutant (12.4Å) | deleterious effect (0.23) thermodynamic stability - destabilising thermal stability - destabilising |
| ATP8B1 | . | R628Q | Nucleotide binding domain | stabilising, favourable torsion angle of Gln residue,  ∆∆G 0.28kcal/mol | destabilising -0.805kcal/mol | destabilising -0.76kcal/mol | destabilising -0.69kcal/mol | decrease in molecule flexibility  ΔΔSVib ENCoM:  -0.88kcal/mol-1/K-1 | WT (4.5Å) to Mutant (4.5Å) | neutral effect (-0.18) |
| NR1H4 | . | N358H | Ligand-binding domain | stabilising, favourable torsion angle of His residue,  ∆∆G 0.75kcal/mol | destabilising -0.758kcal/mol | stabilising 0.050kcal/mol | destabilising -0.752kcal/mol | decrease in molecule flexibility  ΔΔSVib ENCoM:  -0.157kcal/mol-1/K-1 | WT (3.1Å) to Mutant (3.3Å) | neutral effect (-0.67) |
| NR1H4 | . | M173T | Unresolved 3D structure region |  |  |  |  |  |  |  |
